# The Performance of Digital Technologies for Measuring Tuberculosis Medication Adherence: A Systematic Review

**DOI:** 10.1101/2024.05.24.24307886

**Authors:** Miranda Zary, Mona Salaheldin Mohamed, Cedric Kafie, Chimweta Ian Chilala, Shruti Bahukudumbi, Nicola Foster, Genevieve Gore, Katherine Fielding, Ramnath Subbaraman, Kevin Schwartzman

**Affiliations:** McGill International Tuberculosis Centre; Research Institute of the McGill University Health Centre, Montréal, Canada; TB Centre, London School of Hygiene and Tropical Medicine, London, UK; Department of Public Health and Community Medicine, Tufts University School of Medicine, Boston, USA; Schulich Library of Physical Sciences, Life Sciences, and Engineering; McGill University, Montréal, Canada; Division of Geographic Medicine and Infectious Diseases, Tufts Medical Center, Boston, USA

**Keywords:** digital adherence technology, systematic review, accuracy, tuberculosis, performance, treatment observation, medication adherence

## Abstract

**Introduction:** Digital adherence technologies (DATs), such as phone-based technologies, and digital pillboxes, can provide more person-centric approaches to support tuberculosis (TB) medication adherence. We synthesized evidence addressing the performance of DATs for measuring tuberculosis medication adherence.

**Methods:** We conducted a systematic review (PROSPERO - CRD42022313526) which identified relevant published literature from January 2000 through April 2023 in five databases, and pertinent preprints. Studies reporting quantitative data on the performance of DATs for measuring adherence to medications for TB disease or infection, against a reference standard, with at least 20 participants using the DAT were included. Study characteristics and performance outcomes (e.g., sensitivity, specificity, positive and negative predictive values) were extracted. Article quality was assessed using the QUADAS-2 tool for diagnostic accuracy studies.

**Results:** Of 5692 studies initially identified by our systematic search, 13 met our inclusion criteria. These studies addressed the performance of medication sleeves with phone calls [branded as “99DDOTS”; N=4], digital pillboxes [N=5], ingestible sensors [N=2], artificial intelligence-based video observed therapy [N=1], and multifunctional mobile applications [N=1]. All but one involved persons with TB disease. For medication sleeves with phone calls, compared to urine analysis, reported sensitivity and specificity was 70-94% and 0-61%, respectively. For digital pillboxes, compared to pill count, reported sensitivity and specificity was 25-99% and 69-100%, respectively. For ingestible sensors, the sensitivity of dose detection was ≥95% in comparison to directly observed ingestion. Participant selection was the most frequent potential source of bias across articles.

**Conclusion:** Limited available data suggest suboptimal and variable performance of DATs for dose monitoring, with significant evidence gaps, notably in real-world programmatic settings. Future research should aim to improve understanding of the relationships of specific technologies, settings, user characteristics, and user engagement with DAT performance, and should measure and report performance in a more standardized manner.

**KEY MESSAGES:** *What is already known on this topic:* Several cohort studies have suggested that digital adherence technologies (DATs) can both underestimate and overestimate medication ingestion among persons treated for tuberculosis. No previous review has synthesized available evidence in this regard.

*What this study adds:* Reports of DAT (medication sleeves with phone calls, digital pillboxes) implementation in real-world treatment settings consistently indicate suboptimal performance for measuring medication adherence. However, available evidence is limited in scope and quality.

*How this study might affect research, practice, or policy:* Suboptimal dose reporting from DATs potentially compromises their effectiveness, and program efficiency. Future clinical practice will be strengthened by rigorous technology evaluations that reflect more consistent use of reference standards, and clearer benchmarks for medication adherence.

## INTRODUCTION

Tuberculosis (TB) disease requires treatment with medication regimens involving varying pill burdens lasting at least 4 months. Adherence to these treatment regimens—which involve daily medication intake—is crucial, as non-adherence can lead to treatment failure, relapse, development of drug resistance, ongoing TB transmission, and death.(1,2) Treatment for TB infection is between 1 and 9 months. Adherence to treatment for TB infection is essential to reduce the risk of developing TB disease but is often difficult to achieve in routine care, particularly since treated persons are asymptomatic.(3,4) Directly observed therapy (DOT) has been recommended for TB treatment support.(5) DOT involves healthcare workers, or community workers, watching up to 100% of prescribed medication doses.(6) However, DOT is logistically challenging, expensive, potentially intrusive, raises ethical concerns, and can have limited or varying effectiveness for improving treatment outcomes.(6–8)

Digital adherence technologies (DATs) have been increasingly studied and used in routine care as an alternative or adjunctive approach for supporting TB treatment. DATs include mobile communication and other innovations that can remind people with TB to take their medication, digitally observe doses taken, compile dosing histories, triage people who may be at higher risk for unfavorable treatment outcomes, and enable differentiated (i.e., intensified or individualized) care.(2) DATs include a range of technologies that may facilitate more person-centered approaches for monitoring adherence, potentially improving treatment outcomes.(2)

For example, one of the most widely used DATs, branded as 99DOTS, involves wrapping a paper sleeve over a medication blister pack. Dispensation of a medication dose then reveals a hidden phone number. By calling this number, the person with TB can report dose ingestion, creating a digital dosing history that allows early identification by healthcare providers of people who may be nonadherent.(9–12) However, people with TB may call the phone number on the medication sleeve without taking their doses (i.e., over-reporting adherence) or take medication doses without calling (i.e., under-reporting adherence), which may hinder the ability to identify people experiencing nonadherence. Other DATs that have been used in routine care by TB programs (13,14) —such as two-way short messaging service (SMS), digital pillboxes, or video-supported therapy—similarly involve an SMS response, pillbox opening, or remote visualization of dosing by video, respectively, serving as a proxy for a treatment dose taken. All may face challenges related to over-reporting or under-reporting of adherence.

An initial systematic review examining DATs for TB treatment support was published in 2018. However, since 2018—particularly with the COVID-19 pandemic—interest in and experience with these technologies have expanded substantially.(15) While other reviews have been published subsequently (2,16,17), none has examined the performance of DATs for measuring TB medication adherence, which is crucial for healthcare providers to identify people with TB who may be experiencing nonadherence, so they can be given intensified or individualized support. If they do not yield accurate assessments of adherence, any resulting actionable information is of questionable value, resulting in limited public health impact.(2) The present systematic review synthesizes evidence on the performance of digital adherence technologies for measuring TB medication adherence, among persons treated for tuberculosis disease and infection.

## METHODS

### Design

Our systematic review protocol was registered in PROSPERO, the International Prospective Register of Systematic Reviews (CRD42022313526).(18) This review follows the Preferred Reporting Items for Systematic Reviews and Meta-Analyses (PRISMA) guidelines.

### Search and screening strategy

The search for relevant literature was conducted on April 28^th^, 2023 (updated from April 14, 2022) in MEDLINE/Ovid, Embase, CENTRAL, CINAHL, and Web of Science Core Collection, Europe PMC preprints (including MedRxiv) and clinicaltrials.gov from January 1, 2000, to April 28, 2023. Key search concepts included TB (disease or infection), digital technologies (such as mobile phone, smartphone, video observation, digital pillboxes, and text messaging), and accuracy (such as sensitivity, specificity, and area under the curve). The complete search strategy can be found in Table S1 of the supplemental appendix. The database searches were conducted by a health librarian (GG). Separately, we hand-searched the Union World Conference on Lung Health for relevant abstracts on DATs and performance from 2004 to 2022 inclusively. There were no language restrictions.

### Inclusion/Exclusion Criteria

Studies were included if they reported a quantitative outcome addressing the performance of DATs for measuring TB medication adherence (i.e., any of sensitivity, specificity, positive predictive value [PPV], negative predictive value [NPV], area under the receiver operating characteristic curve [AUC], likelihood ratio, accuracy, or agreement). These are defined below (Table 1). Articles were included if the number of participants using the DAT was at least 20, and the study design included the comparison of adherence reports generated by a DAT with a reference standard such as urine drug metabolite testing, pill count, direct observation of medication ingestion, or other such information. DAT interventions included but were not limited to smartphone-based technologies such as phone-based dosing records, SMS or video-supported treatment, digital pillboxes, and ingestible sensors. We defined a DAT as an intervention with a digital component (which could be part of a multi-component intervention) with the intention to measure and promote treatment adherence and/or reduce missed visits and/or reduce losses to follow-up. Examples of DATs included and excluded with this definition can be found in Table S2.

**Table 1:** Definitions of parameters used to describe the performance of DATs for measuring tuberculosis medication adherence. Parameters are defined against a reference standard and by unit of assessment.

| Performance Parameter | Definition – Dose | Definition - Person |
| --- | --- | --- |
| Sensitivity | The percentage of doses that were classified as taken by the DAT, among those that were taken by the reference standard. | The percentage of participants who were classified as adherent by the DAT, among those who were adherent by the reference standard. |
| Specificity | The percentage of doses that were classified as not taken by the DAT, among those that were not taken by the reference standard. | The percentage of participants who were classified as nonadherent by the DAT, among those who were non-adherent by the reference standard. |
| Positive predictive value | Likelihood of a dose being taken by the reference standard among those classified as taken by the DAT. | Likelihood of a participant being adherent by the reference standard among those classified as being adherent by the DAT. |
| Negative predictive value | Likelihood of a dose not being taken by the reference standard among those classified as not being taken by the DAT. | Likelihood of a participant being non-adherent by the reference standard among those classified as being non-adherent by the DAT. |
| Accuracy | The percentage of doses that were classified as taken or not taken by both the DAT and the reference standard. | The percentage of participants who were classified as adherent or non-adherent by both the DAT and the reference standard. |

We included studies of persons treated for TB disease or infection, including subgroups such as people with drug-resistant TB and people with human immunodeficiency virus (HIV). Eligible study designs included all observational studies except case-control studies.(19) We excluded reports if they did not, in fact, involve a DAT (Table S2), or if they were reviews, abstracts (other than from the Union World Conference on Lung Health), editorials, commentaries, news articles, or study protocols. Relevant grey literature (such as ministry reports, technical papers, and preprints) was accepted if it met the eligibility criteria. A complete list of the outcomes, population, intervention, and control groups of interest, and all inclusion/exclusion criteria can be found in Table S3.

### Study Selection

After de-duplication using EndNote (Version 20.2.1 – Clarivate, London UK), five reviewers (M.Z., M.S., C.C., S.B., C.K., and N.F.) independently screened all titles and abstracts for their relevance to DATs for TB treatment support, supported by Rayyan.ai (Rayyan, Cambridge USA).(20,21) Potentially relevant studies underwent independent full-text review by the same reviewers, for eligibility according to the inclusion criteria above. Each screening stage was conducted in duplicate, by two reviewers blinded to each other’s assessment, with conflicts resolved by a senior investigator (K.S., R.S., or K.F.). All references from and citations of each included publication were also screened for inclusion using Google Scholar (Alphabet, Mountain View USA).(22)

### Data extraction

For each included study, data were extracted into a pre-specified Excel (Microsoft, Redmond USA) template by two independent reviewers (M.Z. and M.S., or C.K.) in parallel, and subsequently compared for any discrepancies. Conflicts were resolved by consensus and discussion with a third reviewer (K.S.) when necessary. Extracted data included study characteristics e.g., study design, study setting (i.e., geographic location; inpatient or outpatient), participant characteristics, DAT used, reference standard used, the approaches to classifying adherence for both the DAT and reference standard (e.g., time frame and frequency of adherence assessment), and any important gaps noted by the reviewers. For each study, we extracted all reported performance parameters e.g., true positives (TP), true negatives (TN), false positives (FP), false negatives (FN), sensitivity, specificity, PPV, NPV, accuracy, and AUC.

### Performance parameter definitions

The performance of digital technologies for measuring TB medication adherence can include an assessment of the technical performance or functioning of the DAT, or the performance of the DAT for detecting human behaviour. Technical performance is assessed in controlled conditions to determine whether the designated hardware or software of the DAT can adequately detect doses known to be taken. The performance of a DAT for detecting human behaviour is assessed in real-world conditions to determine its ability to detect a person’s adherence to their medication during their treatment course. In either condition, DAT performance can be assessed per person or per medication dose. Table 1 contains the definitions of the performance parameters used in this review by the unit of assessment: dose vs. person.

### Data synthesis

The extracted data were summarized in tabular form. Pre-specified subgroup analyses were performed where appropriate, addressing specific DATs, TB disease vs. infection, and groups at risk of unfavorable outcomes. Sensitivity and specificity estimates were displayed according to DAT type in forest plots created using RevMan (Version 5.4 – Cochrane, London UK).(23) We calculated pertinent performance parameters that were not directly reported when these could be derived from the underlying data. For parameters that were reported without binomial 95% confidence intervals (CI), they were calculated, when possible, using the Clopper-Pearson Exact Method in R (Version 4.1.2 – GNU Project).(24) This method was also used for articles that had repeated observations per individual, and therefore does not account for within-individual clustering.

Publication bias was assessed qualitatively using Deek’s test for diagnostic accuracy studies.(25) We created a funnel plot of the association between the diagnostic odds ratio (DOR) and the effective sample size (ESS) of each study.(25) Quantitative assessment for publication bias using the associated regression test of asymmetry could not be performed, given the small number of included studies.

### Quality assessment

The included articles were assessed for risk of bias and applicability concerns using the Quality Assessment of Diagnostic Accuracy Studies 2 (QUADAS-2) tool for primary diagnostic accuracy studies.(26) This tool addresses participant selection, the performance of the index test, the performance of the reference test, and the flow and timing of the tests. For each feature, three scores could be used: low, unclear, or high. Two reviewers (M.Z. and M.S., or C.K.) independently assessed each study for quality. Any conflicts were resolved by consensus and discussion with a third reviewer (K.S.) when necessary. Quality assessment results are displayed in graphic and summary format using RevMan (Version 5.4 – Cochrane, London UK).(23)

### GRADE assessment

Finally, we rated the robustness of evidence using the Grading of Recommendations, Assessment, Development, and Evaluation (GRADE) approach for diagnostic tests and strategies.(27) The GRADEpro Guideline Development Tool (McMaster University, Canada & Evidence Price, EU) was used for recommendations.(28) The risk of bias and indirectness was assessed using the relevant elements of the QUADAS-2 checklist. Inconsistency and imprecision were assessed based on outcome variability and confidence interval ranges across articles.

### Patient and public involvement

Patients and the public were not specifically involved in the design, conduct, reporting, or dissemination plans of our research.

## RESULTS

### Study selection

Figure 1 illustrates the PRISMA 2020 flow chart, beginning with the records identified by our search. After de-duplication, there were 5692 records identified, of which 5290 titles and abstracts were not relevant to TB and DATs. Of the remaining 402 reports which underwent full-text review, nine met our inclusion criteria. Four additional reports were identified by hand-searching, for a total of 13 included reports.

**Figure 1:**
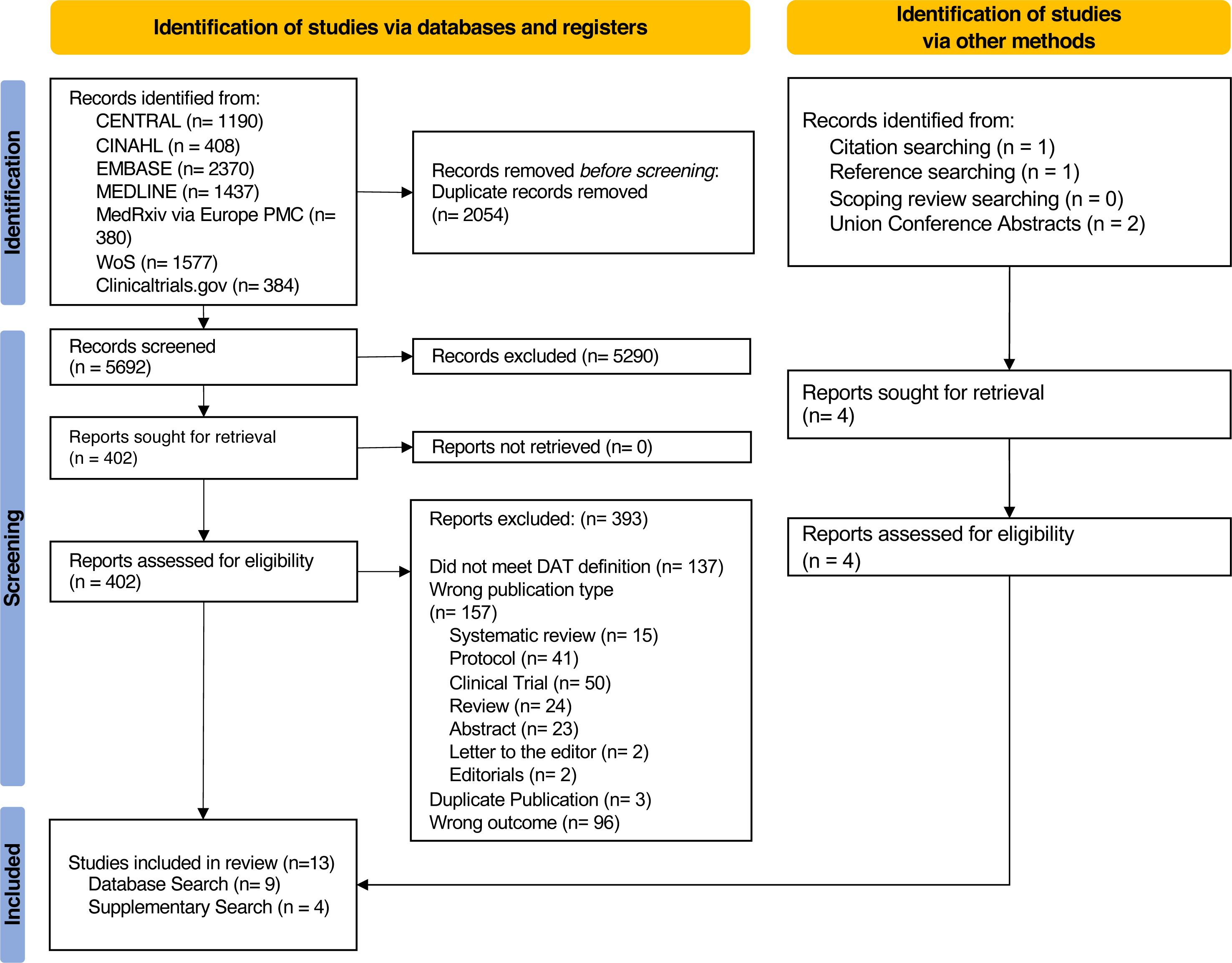
PRISMA 2020 Flow Diagram of studies identified and included. Studies were identified via databases and registries, and via other methods related to the performance of DATs for measuring TB medication adherence. Other methods included searching the references and citations of included studies identified from databases and registries, searching the references and citations of the studies included our linked systematic reviews, and searching the abstracts of the Union World Conference of Lung Health and Tuberculosis from 2004 to 2022. Database and registry searches were conducted on April 28, 2023 from January 1, 2000 to April 28, 2023.

### Overview of studies

Of the 13 reports included, 4 involved medication sleeves with phone calls, 5 involved digital pillboxes, 2 involved ingestible sensors, and 1 each involved a mobile application and an artificial intelligence (AI)-based technology for viewing videos of medication ingestion (known as video observed therapy [VOT]). The study characteristics of the included articles can be found in Table 2. Ingestible sensors, VOT (AI result), and the mobile application were assessed for their technical performance under controlled conditions, while medication sleeves with phone calls and digital pillboxes were assessed under real-world conditions for their ability to detect adherence during TB treatment. Detailed descriptions of the DATs and reference standards used for performance assessment for all articles can be found in Table S4. Briefly, included articles used either isoniazid (INH) urine metabolite tests, rifampicin (RIF) urine colour tests, pill counts, DOT, or healthcare provider-based adherence reports to assess the performance of the DAT (Table 2). 12 articles reported on persons treated for TB disease, while one considered the performance of a digital pillbox in persons treated for TB infection. Two studies compared pill counts and urine drug tests with the digital pillbox as their reference standard; we back-calculated performance parameters for the pillbox compared with pill counts and urine tests using the primary data they reported (Table 2). One study investigating digital pillboxes (Huan et al. 2012) was translated from Simplified Chinese to English using Google Translate (Alphabet, Mountain View USA).(29)

**Table 2:**
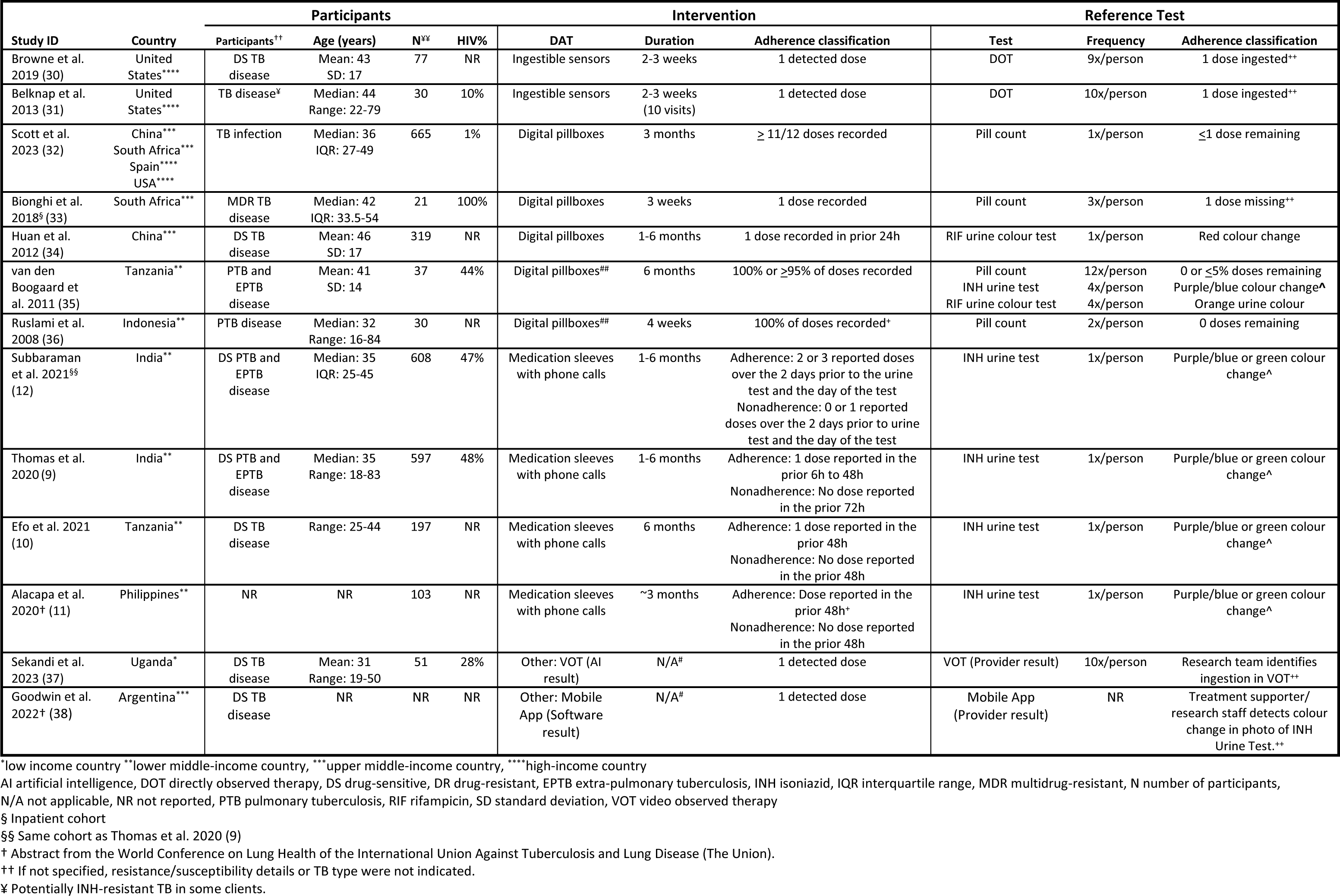
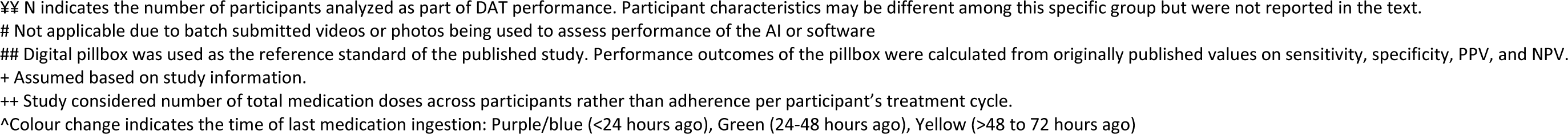
Characteristics of included studies assessing the performance DATs for measuring TB medication adherence.

| Study ID | Country | Participants |  |  |  | Intervention |  |  | Reference Test |  |  |
| --- | --- | --- | --- | --- | --- | --- | --- | --- | --- | --- | --- |
|  |  | Participants<sup>††</sup> | Age (years) | N<sup>‡‡</sup> | HIV% | DAT | Duration | Adherence classification | Test | Frequency | Adherence classification |
| Browne et al. 2019 (30) | United States**** | DS TB disease | Mean: 43<br>SD: 17 | 77 | NR | Ingestible sensors | 2-3 weeks | 1 detected dose | DOT | 9x/person | 1 dose ingested<sup>††</sup> |
| Belknap et al. 2013 (31) | United States**** | TB disease<sup>‡</sup> | Median: 44<br>Range: 22-79 | 30 | 10% | Ingestible sensors | 2-3 weeks<br>(10 visits) | 1 detected dose | DOT | 10x/person | 1 dose ingested<sup>††</sup> |
| Scott et al. 2023 (32) | China***<br>South Africa***<br>Spain****<br>USA**** | TB infection | Median: 36<br>IQR: 27-49 | 665 | 1% | Digital pillboxes | 3 months | ≥ 11/12 doses recorded | Pill count | 1x/person | ≤1 dose remaining |
| Bionghi et al. 2018<sup>§</sup> (33) | South Africa*** | MDR TB disease | Median: 42<br>IQR: 33.5-54 | 21 | 100% | Digital pillboxes | 3 weeks | 1 dose recorded | Pill count | 3x/person | 1 dose missing<sup>††</sup> |
| Huan et al. 2012 (34) | China*** | DS TB disease | Mean: 46<br>SD: 17 | 319 | NR | Digital pillboxes | 1-6 months | 1 dose recorded in prior 24h | RIF urine colour test | 1x/person | Red colour change |
| van den Boogaard et al. 2011 (35) | Tanzania** | PTB and EPTB disease | Mean: 41<br>SD: 14 | 37 | 44% | Digital pillboxes<sup>##</sup> | 6 months | 100% or ≥95% of doses recorded | Pill count<br>INH urine test<br>RIF urine colour test | 12x/person<br>4x/person<br>4x/person | 0 or ≤5% doses remaining<br>Purple/blue colour change<sup>^</sup><br>Orange urine colour |
| Ruslami et al. 2008 (36) | Indonesia** | PTB disease | Median: 32<br>Range: 16-84 | 30 | NR | Digital pillboxes<sup>##</sup> | 4 weeks | 100% of doses recorded<sup>+</sup> | Pill count | 2x/person | 0 doses remaining |
| Subbaraman et al. 2021<sup>§§</sup> (12) | India** | DS PTB and EPTB disease | Median: 35<br>IQR: 25-45 | 608 | 47% | Medication sleeves with phone calls | 1-6 months | Adherence: 2 or 3 reported doses over the 2 days prior to the urine test and the day of the test<br>Nonadherence: 0 or 1 reported doses over the 2 days prior to urine test and the day of the test | INH urine test | 1x/person | Purple/blue or green colour change<sup>^</sup> |
| Thomas et al. 2020 (9) | India** | DS PTB and EPTB disease | Median: 35<br>Range: 18-83 | 597 | 48% | Medication sleeves with phone calls | 1-6 months | Adherence: 1 dose reported in the prior 6h to 48h<br>Nonadherence: No dose reported in the prior 72h | INH urine test | 1x/person | Purple/blue or green colour change<sup>^</sup> |
| Efo et al. 2021 (10) | Tanzania** | DS TB disease | Range: 25-44 | 197 | NR | Medication sleeves with phone calls | 6 months | Adherence: 1 dose reported in the prior 48h<br>Nonadherence: No dose reported in the prior 48h | INH urine test | 1x/person | Purple/blue or green colour change<sup>^</sup> |
| Alacapa et al. 2020<sup>†</sup> (11) | Philippines** | NR | NR | 103 | NR | Medication sleeves with phone calls | ~3 months | Adherence: Dose reported in the prior 48h<sup>+</sup><br>Nonadherence: No dose reported in the prior 48h | INH urine test | 1x/person | Purple/blue or green colour change<sup>^</sup> |
| Sekandi et al. 2023 (37) | Uganda* | DS TB disease | Mean: 31<br>Range: 19-50 | 51 | 28% | Other: VOT (AI result) | N/A<sup>‡</sup> | 1 detected dose | VOT (Provider result) | 10x/person | Research team identifies ingestion in VOT<sup>††</sup> |
| Goodwin et al. 2022<sup>†</sup> (38) | Argentina*** | DS TB disease | NR | NR | NR | Other: Mobile App (Software result) | N/A<sup>‡</sup> | 1 detected dose | Mobile App (Provider result) | NR | Treatment supporter/ research staff detects colour change in photo of INH Urine Test.<sup>††</sup> |

\*low income country \*\*lower middle-income country, \*\*\*upper middle-income country, \*\*\*\*high-income country
AI artificial intelligence, DOT directly observed therapy, DS drug-sensitive, DR drug-resistant, EPTB extra-pulmonary tuberculosis, INH isoniazid, IQR interquartile range, MDR multidrug-resistant, N number of participants,
N/A not applicable, NR not reported, PTB pulmonary tuberculosis, RIF rifampicin, SD standard deviation, VOT video observed therapy
§ Inpatient cohort
§§ Same cohort as Thomas et al. 2020 (9)
† Abstract from the World Conference on Lung Health of the International Union Against Tuberculosis and Lung Disease (The Union).
†† If not specified, resistance/susceptibility details or TB type were not indicated.
‡ Potentially INH-resistant TB in some clients.
¥¥ N indicates the number of participants analyzed as part of DAT performance. Participant characteristics may be different among this specific group but were not reported in the text.

Assessment of adherence by the reference standard was conducted anywhere from once to twelve times over participants’ treatment and the duration of DAT use ranged from two weeks to six months (Table 2). For two studies, authors were contacted with requests to add missing data, but they did not reply. Six of the full articles reported some but not all applicable performance parameters (sensitivity, specificity, PPV, NPV, accuracy; Table S5). While two studies estimated area under the receiver operating characteristic (ROC) curve (Table S5), none provided ROC curves, likelihood ratios, or estimated agreement. When possible, we calculated missing performance parameters from the data provided (Table S5). A meta-analysis pooling performance estimates was not conducted given the small number and marked heterogeneity of the studies retrieved. Only one (30) of five studies (30,31,33,37,38) that analyzed repeated observations of individuals accounted for clustering in their primary analysis. Summary ROC curves to demonstrate joint sensitivity and specificity were not created due to the small number of studies per DAT.

### Performance of DATs under controlled conditions

*Ingestible sensors.* Two reports from the US addressed the performance of ingestible sensors for measuring TB medication adherence in controlled conditions (Table 2).(30,31) This involved determining if an on-body wearable sensor could detect the ingestible sensor placed on the medication, following its ingestion which was directly observed. The percentage of doses that were correctly classified as taken by the sensor’s technology (sensitivity) was at least 95% across studies (Figure 2a, Table S6). Only sensitivity could be estimated from these studies.

**Figure 2:**
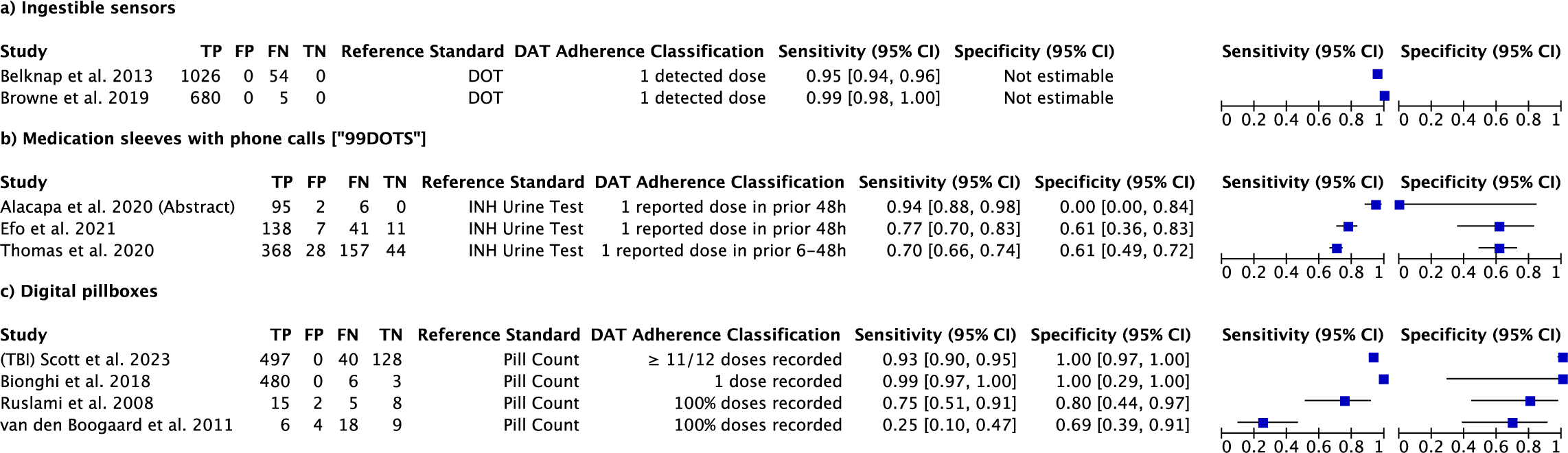
Forest plot of the sensitivity and specificity of a) ingestible sensors, b) Medication sleeves with phone calls [“99DOTS”], and c) Digital pillboxes. a) DOT was used as the reference standard. Sensitivity was calculated using the reported positive detection accuracy of each study (# of ingestible sensors detected/# of ingestible sensors ingested). Specificity was not estimable due to the lack of false positives and true negatives. Browne et al. (30) confidence intervals account for within-individual clustering. b) INH urine test was used as the reference standard. DAT adherence was recorded using “patient-reported doses”, wherein dose reporting relies only on calls made by the person receiving treatment. c) Pill count was used as the reference standard. Results are depicted for persons with tuberculosis disease or infection. Bionghi et al. (33) was an inpatient cohort. Scott et al. (32) used a cohort of persons with TB infection (TBI). In Ruslami et al. (36) and van den Boogaard et al. (35), the digital pillbox was used as the reference standard. Number of true positives, false positives, false negatives, and true negatives were back-calculated from the article’s primary data on pill count sensitivity and specificity, and number of events.

*Other DATs.* Two reports assessed other DATs for their ability to measure TB medication adherence under controlled conditions. In one, a deep convolutional neural network (DCNN) was used to determine whether a participant had ingested medication in a video submitted as part of a VOT intervention.(37) The performance of the artificial intelligence algorithm was compared to the research team’s interpretation of the same submitted video and assessed using 5-fold cross validation, for five types of convolutional neural networks to extract features of the videos. The best performing convolutional neural network had a mean sensitivity of 95% (standard deviation, SD 2.6) but a specificity of 55% (SD 6.5) (Table S6).

In the other report., a mobile application involved the participant’s uploading a photo of their INH urine test, and associated software then analyzed the images to determine whether or not they indicated the presence of INH metabolites.(38) The output from this reader software was compared to treatment supporter and research staff interpretation of the same urine test images. The sensitivity was 81% and 86% when compared to the treatment supporter and research staff interpretations, respectively, while the percentage of doses correctly classified by the technology as not taken (specificity) was 95% and 91% (Table S6).

### Performance of DATs under real-world conditions

*Medication sleeves with phone calls [“99DOTS”]*. Three studies investigated the performance of medication sleeves with phone calls for measuring TB medication adherence, compared with unannounced INH urine tests either in the clinic or at a home visit (Table 2). They generally classified adherence as any dose reported within 48 hours before the urine test and nonadherence as no reported dose within the previous 48 or 72 hours before the urine test. (9–11) All studies investigating this DAT were conducted in lower middle-income countries. The sensitivity of participants’ dose reports (patient-reported doses) for detecting adherence ranged from 70% to 94%, while the specificity of participants’ dose reports for detecting nonadherence ranged from 0% to 61% (Figure 2b).

Two articles on medication sleeves with phone calls, using data from the same cohort, showed that sensitivity and specificity vary depending on the approach used to classify DAT adherence. For example, when “nonadherence” was defined as a person with TB not reporting any medication dose during the 72 hours prior to INH urine testing (9), the specificity was lower than when “nonadherence” was defined as a person reporting less than either two or three medication doses in the 72 hours prior to the home visit (Table 3).(12)

**Table 3:**
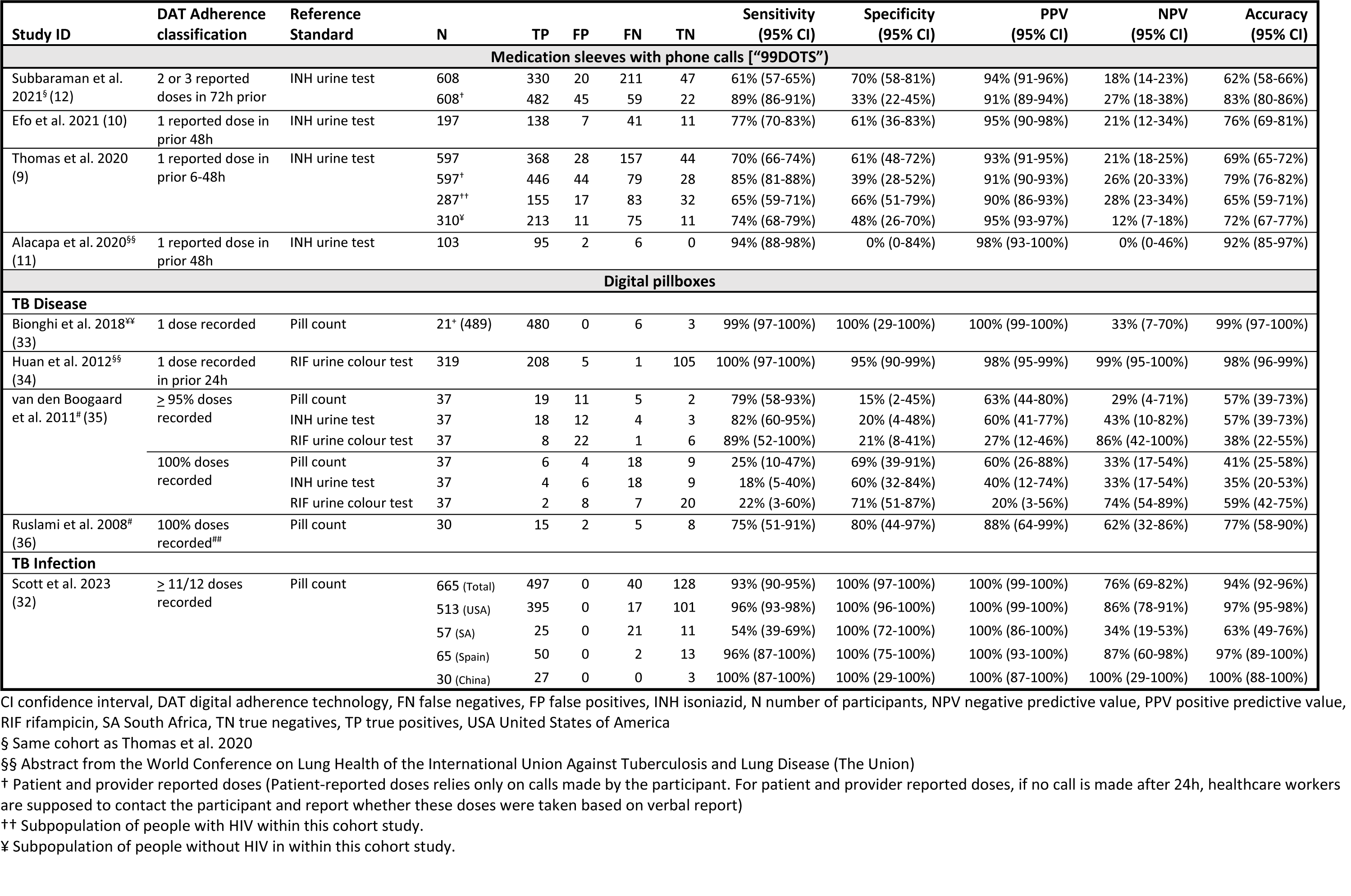
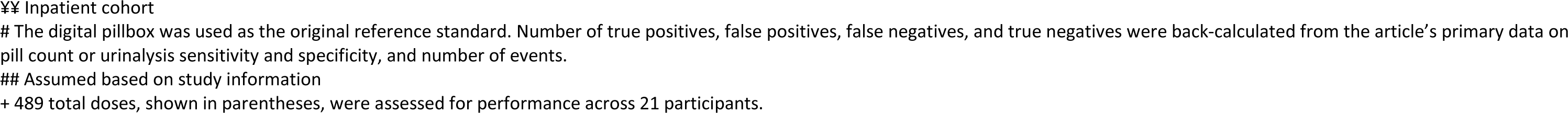
Performance of medication sleeves with phone calls and digital pillboxes under real-world conditions against a reference standard. Digital pillboxes were assessed for persons with tuberculosis disease and tuberculosis infection. If not reported, values and binomial confidence intervals were calculated from other reported performance data (sensitivity, specificity, PPV, NPV, or number of TP, FP, FN, and TN).

| Study ID | DAT Adherence classification | Reference Standard | N | TP | FP | FN | TN | Sensitivity (95% CI) | Specificity (95% CI) | PPV (95% CI) | NPV (95% CI) | Accuracy (95% CI) |
| --- | --- | --- | --- | --- | --- | --- | --- | --- | --- | --- | --- | --- |
| **Medication sleeves with phone calls ["99DOTS"]** |  |  |  |  |  |  |  |  |  |  |  |  |
| Subbaraman et al. 2021<sup>§</sup> (12) | 2 or 3 reported doses in 72h prior | INH urine test | 608 | 330 | 20 | 211 | 47 | 61% (57-65%) | 70% (58-81%) | 94% (91-96%) | 18% (14-23%) | 62% (58-66%) |
|  |  |  | 608<sup>†</sup> | 482 | 45 | 59 | 22 | 89% (86-91%) | 33% (22-45%) | 91% (89-94%) | 27% (18-38%) | 83% (80-86%) |
| Efo et al. 2021 (10) | 1 reported dose in prior 48h | INH urine test | 197 | 138 | 7 | 41 | 11 | 77% (70-83%) | 61% (36-83%) | 95% (90-98%) | 21% (12-34%) | 76% (69-81%) |
| Thomas et al. 2020 (9) | 1 reported dose in prior 6-48h | INH urine test | 597 | 368 | 28 | 157 | 44 | 70% (66-74%) | 61% (48-72%) | 93% (91-95%) | 21% (18-25%) | 69% (65-72%) |
|  |  |  | 597<sup>†</sup> | 446 | 44 | 79 | 28 | 85% (81-88%) | 39% (28-52%) | 91% (90-93%) | 26% (20-33%) | 79% (76-82%) |
|  |  |  | 287<sup>††</sup> | 155 | 17 | 83 | 32 | 65% (59-71%) | 66% (51-79%) | 90% (86-93%) | 28% (23-34%) | 65% (59-71%) |
|  |  |  | 310<sup>‡</sup> | 213 | 11 | 75 | 11 | 74% (68-79%) | 48% (26-70%) | 95% (93-97%) | 12% (7-18%) | 72% (67-77%) |
| Alacapa et al. 2020<sup>§§</sup> (11) | 1 reported dose in prior 48h | INH urine test | 103 | 95 | 2 | 6 | 0 | 94% (88-98%) | 0% (0-84%) | 98% (93-100%) | 0% (0-46%) | 92% (85-97%) |
| **Digital pillboxes** |  |  |  |  |  |  |  |  |  |  |  |  |
| **TB Disease** |  |  |  |  |  |  |  |  |  |  |  |  |
| Bionghi et al. 2018<sup>¶¶</sup> (33) | 1 dose recorded | Pill count | 21<sup>†</sup> (489) | 480 | 0 | 6 | 3 | 99% (97-100%) | 100% (29-100%) | 100% (99-100%) | 33% (7-70%) | 99% (97-100%) |
| Huan et al. 2012<sup>§§</sup> (34) | 1 dose recorded in prior 24h | RIF urine colour test | 319 | 208 | 5 | 1 | 105 | 100% (97-100%) | 95% (90-99%) | 98% (95-99%) | 99% (95-100%) | 98% (96-99%) |
| van den Boogaard et al. 2011<sup>†</sup> (35) | ≥ 95% doses recorded | Pill count | 37 | 19 | 11 | 5 | 2 | 79% (58-93%) | 15% (2-45%) | 63% (44-80%) | 29% (4-71%) | 57% (39-73%) |
|  |  | INH urine test | 37 | 18 | 12 | 4 | 3 | 82% (60-95%) | 20% (4-48%) | 60% (41-77%) | 43% (10-82%) | 57% (39-73%) |
|  |  | RIF urine colour test | 37 | 8 | 22 | 1 | 6 | 89% (52-100%) | 21% (8-41%) | 27% (12-46%) | 86% (42-100%) | 38% (22-55%) |
|  | 100% doses recorded | Pill count | 37 | 6 | 4 | 18 | 9 | 25% (10-47%) | 69% (39-91%) | 60% (26-88%) | 33% (17-54%) | 41% (25-58%) |
|  |  | INH urine test | 37 | 4 | 6 | 18 | 9 | 18% (5-40%) | 60% (32-84%) | 40% (12-74%) | 33% (17-54%) | 35% (20-53%) |
|  |  | RIF urine colour test | 37 | 2 | 8 | 7 | 20 | 22% (3-60%) | 71% (51-87%) | 20% (3-56%) | 74% (54-89%) | 59% (42-75%) |
| Ruslami et al. 2008<sup>#</sup> (36) | 100% doses recorded<sup>###</sup> | Pill count | 30 | 15 | 2 | 5 | 8 | 75% (51-91%) | 80% (44-97%) | 88% (64-99%) | 62% (32-86%) | 77% (58-90%) |
| **TB Infection** |  |  |  |  |  |  |  |  |  |  |  |  |
| Scott et al. 2023 (32) | ≥ 11/12 doses recorded | Pill count | 665 (Total) | 497 | 0 | 40 | 128 | 93% (90-95%) | 100% (97-100%) | 100% (99-100%) | 76% (69-82%) | 94% (92-96%) |
|  |  |  | 513 (USA) | 395 | 0 | 17 | 101 | 96% (93-98%) | 100% (96-100%) | 100% (99-100%) | 86% (78-91%) | 97% (95-98%) |
|  |  |  | 57 (SA) | 25 | 0 | 21 | 11 | 54% (39-69%) | 100% (72-100%) | 100% (86-100%) | 34% (19-53%) | 63% (49-76%) |
|  |  |  | 65 (Spain) | 50 | 0 | 2 | 13 | 96% (87-100%) | 100% (75-100%) | 100% (93-100%) | 87% (60-98%) | 97% (89-100%) |
|  |  |  | 30 (China) | 27 | 0 | 0 | 3 | 100% (87-100%) | 100% (29-100%) | 100% (87-100%) | 100% (29-100%) | 100% (88-100%) |

CI confidence interval, DAT digital adherence technology, FN false negatives, FP false positives, INH isoniazid, N number of participants, NPV negative predictive value, PPV positive predictive value, RIF rifampicin, SA South Africa, TN true negatives, TP true positives, USA United States of America
§ Same cohort as Thomas et al. 2020
§§ Abstract from the World Conference on Lung Health of the International Union Against Tuberculosis and Lung Disease (The Union)
† Patient and provider reported doses (Patient-reported doses relies only on calls made by the participant. For patient and provider reported doses, if no call is made after 24h, healthcare workers are supposed to contact the participant and report whether these doses were taken based on verbal report)
†† Subpopulation of people with HIV within this cohort study.
‡ Subpopulation of people without HIV in within this cohort study.
¥¥ Inpatient cohort

The same authors also examined dose verification added by providers, who telephoned persons who had not phoned in their doses on a given day. When provider-reported doses were added, the sensitivity of combined patient-and provider dose reports for detecting adherence increased from 70% [95% CI 66-74%] to 85% [95% CI 81-88%], but specificity for detecting nonadherence decreased from 61% [95% CI 48-72%] to 39% [95% CI 28-52%] (Table 3).(9)

Estimated overall accuracy ranged from 69-92% (Table 3). Nonetheless, the negative predictive value, or likelihood of a participant being non-adherent by the urine test among those classified as being non-adherent by the DAT, was consistently low (range 0-26%). In other words, non-reporting of a dose by a person with TB often did not mean that they had in fact missed it—and instead often signaled limited ongoing engagement with the technology.

*Digital pillboxes*. The five studies reporting on digital pillboxes used varying methods and were conducted in countries with differing income levels (Table 2). Two used the digital pillbox as their reference standard against multiple index tests, so we back-calculated performance estimates from their reported values (Table S5).(35,36) One assessed performance in an inpatient cohort (33), while one focused on TB infection (32). Four reports included pill counts conducted at clinic visits as at least one of their reference standards (32,33,35,36), while one used unannounced urine color testing for rifampin at home visits (34) (Table 2).

When compared against pill count for those with TB disease, three studies reported digital pillbox sensitivities for overall perfect adherence ranging from 25% to 99%, while specificity for detecting nonadherence was higher, ranging from 69% to 100% (Figure 2c).(33,35,36) The report which only used urine color testing for rifampin estimated the sensitivity and specificity of digital pillboxes for dose detection within 24h as 100% (95% CI 97-100%) and 95% (90-99%), respectively (Table 3).(34) In one article, when compared against pill count among persons treated for TB infection, digital pillboxes were very sensitive for adherence (93% [95% CI 90-95%]) and specific for non-adherence (100% [97-100%]) and generally remained so in subgroup analyses involving the various study sites (Table 3).(32)

For digital pillboxes, overall accuracy estimates ranged from 35%-100% (Table 3). As with the medication sleeves, negative predictive values were often poor, meaning that doses not reported by pillbox opening were not necessarily missed.

### Subgroups and special populations

In an inpatient cohort of 21 individuals with both HIV and drug-resistant TB, digital pillboxes had high sensitivity and specificity (>99%) (Table 3).(33) In another study investigating medication sleeves with phone calls, a subgroup analysis of people with HIV treated for TB disease found that specificity was higher (66% [95% CI 51-79%]) among people with HIV (i.e., more nonadherence was correctly detected) than among people without HIV (48% [26-70%]) (Table 3).(9) Several other studies included persons living with HIV among their participants, but did not report results separately for this group (Table 2).(31,35) No report addressed persons younger than 18.

### Quality Assessment

The risk of bias was high or unclear in at least two categories for all but one of the reports assessed. (Figure S1a). Two reports could not be assessed for quality as the limited description of their methods precluded formal assessment of bias.(11,38) Participant selection was a frequent source of potential bias since participants were rarely randomly sampled; in some cases, they had also explicitly consented to take part in DAT implementation projects (10), which could also introduce bias. Timing of DAT performance assessment relative to reference standard measures may also have been an issue. There were fewer concerns about applicability with respect to participant profiles, DATs used, or reference standards (Figure S1b). Details of the quality assessment criteria are provided in Table S7.

### Publication bias

There was visual asymmetry in the funnel plot displaying diagnostic odds ratios and effective sample sizes, indicating possible publication bias (Figure S2). There were too few studies to permit formal quantitative evaluation of publication bias. Additionally, the effective sample sizes do not account for within-individual clustering and are therefore overestimated for these articles (33,37). Several could not be included because of insufficient data to estimate the diagnostic odds ratios.

### GRADE Results

The findings suggested moderate certainty of evidence for the sensitivity of ingestible sensors, because of consistent and precise results, a low risk of bias, but an inability to assess publication bias. The certainty of evidence was very low for medication sleeves with phone calls because of substantial risks of bias, and substantial variation in sensitivity and specificity across articles. Similarly, the certainty of evidence was very low for digital pillboxes. More details on the quality of evidence for each DAT can be found in Table S8-10 and Figure S3. A GRADE assessment was not conducted for the DATs using AI and reader software since these involved only one report each.

## DISCUSSION

Available evidence addressing the performance of DATs for measuring TB medication adherence is limited in scope and quality. Among 13 reports which considered five types of DATs, there was substantial variation in reference standards, definitions of adherence, and adherence classifications, which made it difficult to compare and summarize results. Ingestible sensors showed relatively high sensitivity under experimental conditions in high-income countries, but this technology may be too expensive to scale up in the near future and will face substantial barriers to real world implementation in LMICs with high TB incidence. In general, however, studies of DAT implementation in real world settings (medication sleeves with phone calls, digital pillboxes) found suboptimal performance of these technologies for measuring medication adherence. These findings are concordant with a previous review that assessed the accuracy of electronic monitoring in devices in antiretroviral medication adherence.(39)

To understand the suboptimal performance of DATs during real world implementation, it is important to consider that this reflects the ways in which people undergoing TB treatment do or do not engage with the technology. Engagement may be influenced by technical attributes of the DAT and the motivations, beliefs, social context, and structural barriers faced by people with TB. Suboptimal engagement with DATs—which may result in under-reporting of true medication adherence (i.e., reduced sensitivity, reduced NPV)—can be influenced by limited access to mobile networks and technology, shared cellphone use among multiple household members, and inadequate education on the purpose and appropriate use of the DAT.(40–43) With substantial under-reporting of medication adherence, healthcare providers may have difficulty identifying people who are truly experiencing nonadherence, be unable to routinely reach out to all people with reported nonadherence (e.g., via phone calls or home visits) (44), and may begin to ignore digital adherence data due to its limited performance (42,44).

Conversely, over-reporting of adherence (i.e., reduced specificity, reduced PPVs)—in which people with TB or healthcare providers may report adherence via phone call or pillbox opening despite medication doses not actually ingested—may result from the desire by people with TB to conceal nonadherence or by the desire of healthcare providers to report optimal TB outcomes.(45) Over-reporting of adherence may be more concerning, as healthcare providers and health systems may miss early identification of people who need intensified or personalized support to improve adherence and ensure optimal treatment outcomes.

Several challenges limited our interpretation of DAT performance. The certainty around evidence of performance for DATs under programmatic conditions was very limited. Additionally, the use of different pill taking thresholds to define adherence, and of different windows for dose recording vs. urine testing, adds substantial complexity and makes it even more difficult to summarize the available evidence. One study evaluated digital pillboxes in an inpatient setting, which differs substantially from the settings where DATs would most likely be used and limits the relevance of the resulting data. Another used a method for documenting the presence or absence of rifampin in urine, which was not used in other studies, highlighting the importance of consistent reference standards. More generally, the use of urine tests as reference standards can be problematic, given known gaps in their performance. This includes suboptimal sensitivity (based on witnessed pill ingestion) for doses ingested over 24 hours before testing (46), and suboptimal specificity for doses ingested over 72 hours before testing.(12,47) Similarly, while pill count is a standard method for determining adherence, pills missing during routine pill counts may not have been ingested.

### Strengths and Limitations

To our knowledge, this is the first systematic review focusing specifically on the performance of DATs for measuring TB medication adherence. We used a wide-ranging, pre-specified search strategy across multiple databases, without language restriction and with the potential inclusion of suitable reports from grey literature, including preprints. Our search was developed and conducted by an experienced health sciences librarian. Selection of reports and data extraction were performed rigorously, by two independent reviewers at every step. Whenever possible, we used primary data from each report to estimate any relevant performance parameters that the authors had not reported. We also assessed the quality and robustness of the available evidence.

For logistical reasons, we could not systematically search for and retrieve abstracts other than those presented at the conferences of the International Union Against Tuberculosis and Lung Disease, which is the major venue for public health-oriented TB research. Some reports did not include sufficient data to permit estimation of all performance parameters of interest. Additionally, there may be an underestimation of the variance in instances where clustering was not considered (i.e., for repeated measurements per individual). We did not identify any reports evaluating the performance of doses observed by video against urine tests or pill counts, although video-supported treatment has been used increasingly in high-income countries.(48–50) Finally, given the heterogeneity of study methods, technologies, and results, we did not formally pool or meta-analyze their data. We have instead summarized the key results from each study in tabular and/or graphic format, and provided an overview of the results for each key technology.

### Changes to the protocol

Changes were made to our pre-registered protocol in PROSPERO as follows. We additionally searched Europe PMC for pre-print articles relevant to our review. We did not anticipate the use of DOT, or provider-based DAT decisions as reference standards for quantifying DAT performance, and therefore added them as possible comparators. We also did not anticipate articles that used the DAT as the reference standard. For those reports, we back-calculated the published results to provide us with the performance of the DAT as the index test.

## CONCLUSIONS

Accurate dose reporting is fundamental to the use of digital technologies to aid treatment adherence. Their use as alternatives to traditional direct observation is predicated on the concept that providers and health programs can provide additional support to people who appear to be facing challenges in this regard. Hence suboptimal performance of dose reports from DATs can potentially compromise their effectiveness, as well as program efficiency. If the DAT fails to capture significant non-adherence, this leads to missed opportunities for intervention, and potentially even poorer treatment outcomes. On the other hand, if support and supervision are intensified for people wrongly labeled as poorly adherent by the DAT, this is a waste of limited program resources. The future evidence base will be strengthened by more consistent definitions and cutoffs for adherence, and by more consistent use of one or more reference standards—e.g. validated methods for pill counts, standardized timing and technique for urine tests. Future research should also address the interplay of specific technology, setting, user characteristics, and user engagement with DAT performance. Additional studies examining the performance of asynchronous video observation will also be important, as will further investigation of digital technologies to support newer treatment regimens for TB disease and infection.

## DECLARATIONS

### Ethics approval and consent to participate

Not applicable

### Consent for publication

Not applicable

### Availability of data and materials

The data and materials supporting the conclusions of this article that are not already shared in the manuscript and appendix are available at https://borealisdata.ca/dataverse/mcgill

### Authors’ contributions

All authors contributed to the design of the research and approved the final manuscript. MZ contributed to screening, data extraction and analysis, and drafted the manuscript. MS contributed to the database search, screening, data extraction, and provided systematic review expertise for the manuscript. CK contributed to screening and data extraction. GG contributed to the database search. CC, SB, and NF contributed to screening. KF, RS, and KS provided support in all steps of executing the review and revised the manuscript.

## Supporting information

PRISMA checklist

## Data Availability

All data produced in the present study are contained in the manuscript and supplementary material.

https://borealisdata.ca/dataverse/mcgill

## Acknowledgements

Part of this work was presented in abstract and poster format at the 2023 North America Region Conference of the International Union of Tuberculosis and Lung Disease on Tuberculosis (51).

