## Supplementary tables and figures for "The Performance of Digital Technologies for Measuring Tuberculosis Medication Adherence: A Systematic Review"

#### *Supplementary Materials*

Miranda Zary[1], Mona Salaheldin Mohamed[1], Cedric Kafie[1], Chimweta Ian Chilala[2] , Shruti Bahukudumbi[3], Nicola Foster[2], Genevieve Gore[4], Katherine Fielding[2], Ramnath Subbaraman[3,5], and Kevin Schwartzman[1]

[1]McGill International Tuberculosis Centre; Research Institute of the McGill University Health Centre, Montréal, Canada
[2]TB Centre, London School of Hygiene and Tropical Medicine, London, UK
[3]Department of Public Health and Community Medicine, Tufts University School of Medicine, Boston, USA
[4]Schulich Library of Physical Sciences, Life Sciences, and Engineering; McGill University, Montréal, Canada
[5]Division of Geographic Medicine and Infectious Diseases, Tufts Medical Center, Boston, USA

**Table S1: Search strategy for performance of DATs for measuring TB medication adherence systematic review.**

#### MEDLINE (Ovid)

Ovid MEDLINE(R) ALL <1946 to April 18, 2023>

1 exp tuberculosis/ or mycobacterium tuberculosis/ 228099

2 Antitubercular Agents/ 41251

3 (Tuberculosis or Kochs disease or Phthisis or TB or MTB or MDRTB or XDRTB or DRTB or LTBI or directly observed treatment short course).mp. 300916

4 1 or 2 or 3 [TB CONCEPT] 306343

4 ((antitubercular agents/ or directly observed therapy/ or medication adherence/ or patient compliance/ or "treatment adherence and compliance"/) and technology/) or mobile applications/ or internet/ or cell phone/ or smartphone/ or text messaging/ or computer, handheld/ or telemedicine/ or therapy, computer-assisted/ or medical informatics applications/ 148201

5 (((digital* or electronic* or mobile or wireless* or virtual*) adj2 (adherence or medication monitor* or medication package* or observ*)) or digital technolog* or technology-based or Digital health or eHealth or e health or mhealth or m health or SMS or reminder* or short messag* service* or text messag* or MMS or multimedia messag* or MEMS or (monitor* adj2 (electronic* or sensor* or device*)) or Webcam* or web cam* or smartphone* or smart phone* or web based or health IT or health ICT or ((cell* or mobile) adj1 (device* or health or phone* or technolog*)) or video* or cellphone* or feature phone* or vdot or vmalt or tele* or dat or wot or 99dots or 99 dots or monitoring system* or ingestible sensor* or merm or artificial intelligence or ai or vot or ((digital or smart) adj (pill box* or pillbox*)) or VST).mp. 705421

6 4 or 5 [DAT CONCEPT] 772859

7 13 and 16 [TB CONCEPT AND DAT CONCEPT] 3219

8 limit 17 to yr="2000 -Current" 2791

9 exp "sensitivity and specificity"/ or likelihood functions/ or exp "reproducibility of results"/ 978449

10 (Accura* or Sensitiv* or Specific* or likelihood or Precis* or Valid* or Positive predictive value* or negative predictive value* or Dosage or dosing or dose? or Report or reporting or Area under the curve or AUC or Receiver operator curve* or ROC or Agreement or reliab* or reproduc*).mp. 11893066

11 9 or 10 11980293

12 8 and 11 [TB CONCEPT AND DAT CONCEPT AND ACCURACY CONCEPT] 1437

#### Embase (Ovid)

Embase <1996 to 2023 Week 15>

1 exp tuberculosis/ or mycobacterium tuberculosis/ 204337

2 tuberculostatic agent/ 30987

3 (Tuberculosis or Kochs disease or Phthisis or TB or MTB or MDRTB or XDRTB or DRTB or LTBI or directly observed treatment short course).mp. 246187

4 1 or 2 or 3 [TB CONCEPT] 257637

5 ((tuberculostatic agent/ or directly observed therapy/ or medication compliance/ or patient compliance/) and technology/) or communication technology/ or exp mobile application/ or internet/ or web-based intervention/ or exp mobile phone/ or text messaging/ or personal digital assistant/ or telemedicine/ or exp teleconsultation/ or telemonitoring/ or video consultation/ or computer-assisted therapy/ or computer-assisted drug therapy/ or medical informatics/ 261570

6 (((digital* or electronic* or mobile or wireless* or virtual*) adj2 (adherence or medication monitor* or medication package* or observ*)) or digital technolog* or technology-based or Digital health or eHealth or e health or mhealth or m health or SMS or reminder* or short messag* service* or text messag* or MMS or multimedia messag* or MEMS or (monitor* adj2 (electronic* or sensor* or device*)) or Webcam* or web cam* or smartphone* or smart phone* or web based or health IT or health ICT or ((cell* or mobile) adj1 (device* or health or phone* or technolog*)) or video* or cellphone* or feature phone* or vdot or vmalt or tele* or dat or wot or 99dots or 99 dots or monitoring system* or ingestible sensor* or merm or artificial intelligence or ai or vot or ((digital or smart) adj (pill box* or pillbox*)) or VST).mp. 870210

7 5 or 6 [DAT CONCEPT] 984663

8 4 and 7 [TB CONCEPT AND DAT CONCEPT] 5167

9 limit 8 to yr="2000 -Current" 5020

10 exp "sensitivity and specificity"/ or measurement accuracy/ or reproducibility/ or predictive value/ or predictive validity/ or exp *validity/ 884855

11 (Accura* or Sensitiv* or Specific* or likelihood or Precis* or Valid* or Positive predictive value* or negative predictive value* or Dosage or dosing or dose? or Report or reporting or Area under the curve or AUC or Receiver operator curve* or ROC or Agreement or reliab* or reproduc*).ti,ab,kf. 11272549

12 10 or 11 11451773

13 9 and 12 2370

#### CINAHL (EBSCOhost)

| # | Query | Limiters/Expanders | Last Run Via | Results |
|---|---|---|---|---|
| S13 | S9 AND S12 | Expanders - Apply equivalent subjects Search modes - Boolean/Phrase | Interface - EBSCOhost Research Databases Search Screen - Advanced Search Database - CINAHL Plus with Full Text | 408 |

| | | | | |
|---|---|---|---|---|
| S12 | S10 OR S11 | Expanders - Apply equivalent subjects Search modes - Boolean/Phrase | Interface - EBSCOhost Research Databases Search Screen - Advanced Search Database - CINAHL Plus with Full Text | 2,384,775 |
| S11 | (Accura* OR Sensitiv* OR Specific* OR likelihood OR Precis* OR Valid* OR "Positive predictive value*" OR "negative predictive value*" OR Dosage OR dosing OR dose# OR Report OR reporting OR "Area under the curve" OR AUC OR "Receiver operator curve*" OR ROC OR Agreement OR reliab* OR reproduc*) | Expanders - Apply equivalent subjects Search modes - Boolean/Phrase | Interface - EBSCOhost Research Databases Search Screen - Advanced Search Database - CINAHL Plus with Full Text | 2,384,775 |
| S10 | (MH "Sensitivity and Specificity") OR (MH "Reproducibility of Results") | Expanders - Apply equivalent subjects Search modes - Boolean/Phrase | Interface - EBSCOhost Research Databases Search Screen - Advanced Search Database - CINAHL Plus with Full Text | 147,249 |
| S9 | S4 AND S7 | Limiters - Published Date: 20000101-20231231 Expanders - Apply equivalent subjects Search modes - Boolean/Phrase | Interface - EBSCOhost Research Databases Search Screen - Advanced Search Database - CINAHL Plus with Full Text | 858 |

| | | | | |
|---|---|---|---|---|
| S8 | S4 AND S7 | Expanders - Apply equivalent subjects Search modes - Boolean/Phrase | Interface - EBSCOhost Research Databases Search Screen - Advanced Search Database - CINAHL Plus with Full Text | 917 |
| S7 | S5 OR S6 | Expanders - Apply equivalent subjects Search modes - Boolean/Phrase | Interface - EBSCOhost Research Databases Search Screen - Advanced Search Database - CINAHL Plus with Full Text | 347,116 |
| S6 | (((digital* OR electronic* OR mobile OR wireless* OR virtual*) N2 (adherence OR "medication monitor*" OR "medication package*" OR observ*)) OR "digital technolog*" OR technology-based OR "Digital health" OR eHealth OR "e health" OR mhealth OR "m health" OR SMS OR reminder* OR "short messag* service*" OR "text messag*" OR MMS OR "multimedia messag*" OR MEMS OR (monitor* N2 (electronic* OR sensor* OR device*)) OR Webcam* OR "web cam*" OR smartphone* OR "smart phone*" OR "web based" OR "health IT" OR "health ICT" OR ((cell* OR mobile) N1 (device* OR health OR phone* OR technolog*)) OR video* OR cellphone* OR "feature phone*" OR vdot OR vmalt OR tele* OR dat OR wot OR 99dots OR "99 dots" OR "monitoring system*" OR "ingestible sensor*" OR merm OR "artificial intelligence" OR ai OR vot OR ((digital OR smart) W1 ("pill box*" OR pillbox*)) OR VST) | Expanders - Apply equivalent subjects Search modes - Boolean/Phrase | Interface - EBSCOhost Research Databases Search Screen - Advanced Search Database - CINAHL Plus with Full Text | 278,595 |
| S5 | ( (MH "Antitubercular Agents") OR (MH "Directly Observed Therapy") OR (MH "Medication Compliance") OR (MH "Patient Compliance")) AND (MH "Technology") ) OR (MH "Wireless Communications") OR (MH "Mobile Applications") OR (MH "Internet") OR (MH "Internet-Based | Expanders - Apply equivalent subjects Search modes - Boolean/Phrase | Interface - EBSCOhost Research Databases Search | 133,190 |

| | | | | |
|---|---|---|---|---|
| | Intervention") OR (MH "Cellular Phone+") OR (MH "Text Messaging+") OR (MH "Instant Messaging") OR (MH "Interactive Voice Response Systems") OR (MH "Videoconferencing+") OR (MH "Computers, Hand-Held+") OR (MH "Telehealth+") OR (MH "Digital Technology") OR (MH "Therapy, Computer Assisted") OR (MH "Drug Therapy, Computer Assisted") OR (MH "Medical Informatics") | | Screen - Advanced Search Database - CINAHL Plus with Full Text | |
| S4 | S1 OR S2 OR S3 | Expanders - Apply equivalent subjects Search modes - Boolean/Phrase | Interface - EBSCOhost Research Databases Search Screen - Advanced Search Database - CINAHL Plus with Full Text | 39,412 |
| S3 | (Tuberculosis OR "Kochs disease" OR Phthisis OR TB OR MTB OR MDRTB OR XDRTB OR DRTB OR LTBI OR "directly observed treatment short course") | Expanders - Apply equivalent subjects Search modes - Boolean/Phrase | Interface - EBSCOhost Research Databases Search Screen - Advanced Search Database - CINAHL Plus with Full Text | 39,091 |
| S2 | (MH "Antitubercular Agents") | Expanders - Apply equivalent subjects Search modes - Boolean/Phrase | Interface - EBSCOhost Research Databases Search Screen - Advanced Search Database - CINAHL Plus with Full Text | 4,883 |

| | | | | |
|---|---|---|---|---|
| S1 | (MH "Tuberculosis+") OR (MH "Mycobacterium Tuberculosis") | Expanders - Apply equivalent subjects Search modes - Boolean/Phrase | Interface - EBSCOhost Research Databases Search Screen - Advanced Search Database - CINAHL Plus with Full Text | 26,642 |

#### CENTRAL (Cochrane Library/Wiley)

Search Name:
Date Run: 28/04/2023 21:08:13
Comment:

ID Search Hits
#1 (Tuberculosis or "Kochs disease" or Phthisis or TB or MTB or MDRTB or XDRTB or DRTB or LTBI or "directly observed treatment short course"):ti,ab,kw 9141
#2 (((digital* or electronic* or mobile or wireless* or virtual*) NEAR/2 (adherence or medication NEXT monitor* or medication NEXT package* or observ*)) or digital NEXT technolog* or technology-based or "Digital health" or eHealth or "e health" or mhealth or "m health" or SMS or reminder* or short NEXT messag* NEXT service* or text NEXT messag* or MMS or multimedia NEXT messag* or MEMS or (monitor* NEAR/2 (electronic* or sensor* or device*)) or Webcam* or web NEXT cam* or smartphone* or smart NEXT phone* or "web based" or "health IT" or "health ICT" or ((cell* or mobile) NEXT (device* or health or phone* or technolog*)) or video* or cellphone* or feature NEXT phone* or vdot or vmalt or tele* or dat or wot or 99dots or 99 NEXT dots or monitoring NEXT system* or ingestible NEXT sensor* or merm or artificial NEXT intelligence or ai or vot or ((digital or smart) NEXT (pill box* or pillbox*)) or VST):ti,ab,kw 359219
#3 #1 AND #2 with Publication Year from 2000 to 2023, in Trials 2118
#4 (Accura* or Sensitiv* or Specific* or likelihood or Precis* or Valid* or Positive NEXT predictive NEXT value* or negative NEXT predictive NEXT value* or Dosage or dosing or dose? or Report or reporting or "Area under the curve" or AUC or Receiver NEXT operator NEXT curve* or ROC or Agreement or reliab* or reproduc*):ti,ab,kw 824301
#5 #3 AND #4 1190

#### WOS.SCI, WOS.ISTP, WOS.ESCI (Web of Science Core Collection)

### Web of Science Search Strategy (v0.1)

Search: TS=(Tuberculosis OR "Kochs disease" OR Phthisis OR TB OR MTB OR MDRTB OR XDRTB OR DRTB OR LTBI OR "directly observed treatment short course" )

Editions: WOS.SCI,WOS.ISTP,WOS.ESCI

Date Run: Fri Apr 28 2023 17:49:53 GMT-0400 (Eastern Daylight Time)

Results: 248874

Search: TS=(((digital* OR electronic* OR mobile OR wireless* OR virtual* ) NEAR/2 (adherence OR "medication monitor*" OR "medication package*" OR observ* )) OR "digital technolog*" OR technology-based OR "Digital health" OR eHealth OR "e health" OR mhealth OR "m health" OR SMS OR reminder* OR "short messag* service*" OR "text messag*" OR MMS OR "multimedia messag*" OR MEMS OR (monitor* NEAR/2 (electronic* OR sensor* OR device* )) OR Webcam* OR "web cam*" OR smartphone* OR "smart phone*" OR "web based" OR "health IT" OR "health ICT" OR ((cell* OR mobile ) NEAR/1 (device* OR health OR phone* OR technolog* )) OR video* OR cellphone* OR "feature phone*" OR vdot OR vmalt OR tele* OR dat OR wot OR 99dots OR "99 dots" OR "monitoring system*" OR "ingestible sensor*" OR merm OR "artificial intelligence" OR ai OR vot OR ((digital OR smart ) NEAR/0 ("pill box*" OR pillbox* )) OR VST )

Editions: WOS.SCI,WOS.ISTP,WOS.ESCI

Date Run: Fri Apr 28 2023 17:50:06 GMT-0400 (Eastern Daylight Time)

Results: 1632660

Search: TS=(Accura* OR Sensitiv* OR Specific* OR likelihood OR Precis* OR Valid* OR "Positive predictive value*" OR "negative predictive value*" OR Dosage OR dosing OR dose$ OR Report OR reporting OR "Area under the curve" OR AUC OR "Receiver operator curve*" OR ROC OR Agreement OR reliab* OR reproduc*)

Editions: WOS.SCI,WOS.ISTP,WOS.ESCI

Date Run: Fri Apr 28 2023 17:54:01 GMT-0400 (Eastern Daylight Time)

Results: 18948839

Search: #1 AND #2 AND #3

Editions: WOS.SCI,WOS.ISTP,WOS.ESCI

Date Run: Fri Apr 28 2023 17:54:14 GMT-0400 (Eastern Daylight Time)

Results: 1676

### Database: Web of Science Core Collection

### Entitlements:

- WOS.IC: 1993 to 2023
- WOS.CCR: 1985 to 2023
- WOS.SCI: 1900 to 2023
- WOS.AHCI: 1975 to 2023
- WOS.BHCI: 2005 to 2023
- WOS.BSCI: 2005 to 2023

- WOS.ESCI: 2005 to 2023
- WOS.ISTP: 1990 to 2023
- WOS.SSCI: 1900 to 2023
- WOS.ISSHP: 1990 to 2023

### Searches:

Search: #1 AND #2 AND #3

Editions: WOS.SCI,WOS.ISTP,WOS.ESCI

Timespan: 2000-01-01 to 2023-04-28

Date Run: Fri Apr 28 2023 17:54:40 GMT-0400 (Eastern Daylight Time)

Results: 1577

#### MedRxiv and other preprints via Europe PMC

(TITLE:Tuberculosis OR TITLE:"Kochs disease" OR TITLE:Phthisis OR TITLE:TB OR TITLE:MTB OR TITLE:MDRTB OR TITLE:XDRTB OR TITLE:DRTB OR TITLE:LTBI OR TITLE:"directly observed treatment short course" OR TITLE:"antituberculosis" OR TITLE:"antituberculous") AND (Adher* OR "directly observed" OR Digital OR electronic OR internet OR mobile OR wireless OR virtual OR TITLE:technology OR "technology based" OR tele* OR ehealth OR "e health" OR mhealth OR "m health" OR SMS OR reminder* OR messaging OR message* OR MEMS OR web OR webcam* OR smartphone* OR "health IT" OR "health ICT" OR video* OR cellphone* OR phone* OR vdot OR vmalt OR dat OR wot OR 99dots OR "99 dots" OR "monitoring system" OR "monitoring systems" OR "ingestible sensor" OR "ingestible sensors" OR merm OR "artificial intelligence" OR AI OR VOT OR "smart pillbox" OR "smart pill box" OR VST OR "computer assisted") AND (SRC:PPR)

**Table S2: Definition of a digital adherence technology, with examples for inclusion and exclusion.**

| | |
|---|---|
| Intervention | • A digital component (which could be part of a multi-component intervention)<br>• with the intention to measure or promote treatment adherence and/or reducing missed visits and/or reducing LTFU (and thereby improving successful treatment outcomes)<br><br>The intervention can be patient-facing only, provider-facing only, or patient and provider-facing. |
| *Examples of "digital adherence component" (though not limited to):* | • SMS daily reminders (1- or 2-way) to patient to take treatment.<br>• SMS reminders (1- or 2-way) to patient to attend dispensing visit/routine follow-up appointment.<br>• Automated phone calls to remind patient for visit/daily dose.<br>• Smart pillbox daily reminders to patient to take treatment.<br>• Smart pillbox reminders to patient attend dispensing visit/routine follow-up appointment.<br>• Chatbot/telemedicine accessed by TB patients that provides information about "treatment adherence".<br>• Automated feedback (such as electronic health record) to alert HCW to a missed visit (e.g., dispensing, routine follow-up appt for patients on TB/TBI treatment) by a patient with an intended action of "promote treatment adherence and/or reducing LTFU".<br>• Digital calendar generated (from electronic health record/smart pill box) showing missed visits/doses by patient at consultation with HCW.<br>• Telehealth – video-call initiated by HCW.<br>• WhatsApp group (with patients/HCWs)- educational messages sent etc.<br>• Specialist Healthcare provider Apps (– not automated) – used by patient and HCW.<br>• Electronic pillbox (or suchlike) used to measure adherence only (i.e., not used to promote adherence/improving outcomes)<br><br>These need to be used *in conjunction* with the intention to measure or promote treatment adherence and/or reducing missed visits and/or reducing LTFU (and thereby improving successful treatment outcomes). |
| *Examples to exclude (not limited to)* | • Electronic health record to document (monitor/record/summarise) visit attendance only - with an NO intended action of "measure or promote treatment adherence and/or reducing LTFU".<br>• Mobile technology to collect data (TB register etc..) with no feedback loop except for specific examples of technology used to measure.<br>• Non-automated "routine telephone calls" to patient.<br><br>Phone calls or SMS or WhatsApp (by human) & not automated - with no other digital component ("older-style"). |

**Table S3: PICO Definitions and Inclusion/Exclusion Criteria pre-specified in the PROSPERO systematic review registration.** (1)

| | |
|---|---|
| Participants/population | • Individuals diagnosed and treated for TB infection.<br>• Individuals treated for active TB disease, including those at risk of unfavorable outcomes (e.g., those with drug-resistant TB, persons living with HIV, children). |
| Intervention/Exposure | • DATs which support monitoring TB medication adherence, include reminding patients to take their medications, facilitating digital observation of pill-taking, compiling patient dosing histories and categorizing patients based on their level of adherence, to facilitate patient-centric approaches for monitoring TB medication adherence. See Figure S1.<br>• DATs include and are not limited to smartphone-based technologies such as phone-based dosing records, SMS or Video-supported treatment, digital pillboxes, and ingestible sensors. See Figure S1. |
| Comparator/Control | • Studies included will compare the dose reporting by DAT against a reference standard. This reference standard could be measurement of drug/ metabolite levels or their presence in serum or urine, and/or pill count, or directly observed therapy. |
| Inclusion Criteria | Studies will be included if:<br>• They address tuberculosis in humans.<br>• They address at least one digital adherence technology.<br>• They assess accuracy quantitatively.<br>• There will be no language restriction.<br>• Relevant grey literature (such as ministry reports, technical papers, preprints) will be accepted if they meet the eligibility criteria. |
| Exclusion Criteria | Studies will be excluded if:<br>• They are not themed on the use of digital health interventions for tuberculosis treatment support.<br>• If the same study with the same outcomes has been reported by different papers, - the most recent published paper will be included.<br>• For the purposes of our analysis, we will also exclude reports/ papers where only the abstract but not the full text is available. However, during the search process, we will keep a record of abstracts for which the final study has not been published in a peer-reviewed journal to provide insights into potential publication bias. Publication bias occurs when smaller studies or studies with negative findings do not get published, which can contribute to bias in the findings of meta-analyses. If, for example, we identify a large number of abstracts with negative findings that never got published in peer-reviewed journals, this could be indicative of publication bias.<br>• They are letters, editorials, and position papers. o They are review articles.<br>• They are case control studies.<br>• They report purely qualitative information.<br>• They report fewer than 20 persons receiving treatment support with a DAT. |
| Main outcomes | Accuracy of dose reporting will be assessed by comparison of dose recording by DAT against a reference standard.<br>Primary outcomes will be:<br>• Sensitivity of dose reporting by DAT, against the reference standard<br>• Specificity, assessed in the same manner |
| Additional outcomes | Secondary effect measures of accuracy of DAT dose reporting can include:<br>• Area under the curve |

| | |
|---|---|
| | • Receiver operator curve<br>• Positive predictive value<br>• Likelihood ratios<br>• Negative predictive value<br>• Agreement |

**Table S4: (A) DAT and (B) Reference Standard Characteristics of the included studies.**

**(A) DATs**

| Technology | Details | Adherence and nonadherence classification |
|---|---|---|
| Medication sleeves with phone calls | Fixed-dose combination medication blister packs are placed within a custom cardstock that has a series of toll-free phone numbers that are revealed once the person removes the prescribed dose. After ingestion, the person is expected to call the toll-free number to automatically log their adherence for the day. Healthcare providers (HCPs) visualize this adherence record remotely to identify nonadherent people. Branded as '99DOTS' in all studies.<br>Patient-reported doses (default): Person with TB calling to report their dose.<br>Patient/Provider-reported doses (only assessed in Subbaraman et al.(2) and Thomas et al. (3)): Person with TB calling to report their dose; if the person with TB does not call on a given day or days, the provider is supposed to call the person to verify whether doses were taken or not and then report these data in the digital dosing history. | Refer to Table 2 (main text) for the reporting windows for each study.<br>Previous adherence was assessed at the time of random urinalysis.<br><br>*Efo et al.*(4)*:* SMS reminders automatically sent at specific times of the day prompted people to take their medication.<br>*Thomas et al.* (3)*:* People whose record reported doses taken <6 hours and 48-72 hours prior were excluded. |
| Digital pillbox | Electronic medication bottle, pill cap, or pillbox that contains a microelectronic chip that registers every bottle opening or box opening, respectively. Some devices contain a subscriber identity module (SIM) card and use a cellular network to provide real-time measurements of pillbox openings. Branded as 'MEMS' or 'Wisepill'. | Doses recorded as taken (adherence) or not taken (nonadherence) by the device were assessed per participant or across participants. Records were assessed at provider visits during treatment or at the end of treatment.<br><br>*Scott et al.*(5)*:* One arm of the trial also received text message reminders |
| Ingestible sensors | Consists of a 1.0 mm x 1.0 mm ingestible sensor placed on or taken with medication and an on-body wearable sensor. The ingestible sensors are activated by gastric fluids, independent of the acidity level, and communicate unique identifying signatures to the body surface. The on-body sensor counts the number of times each unique signature is received. Data from the patch are transmitted wirelessly (can use Bluetooth) to a secured device and uploaded to a secured, centralized data storage location. Data could be shared with the participant in near-real time and viewed on their mobile device. Also termed "wirelessly observed therapy (WOT)" and "ingestion sensors". | A detected ingested marker (adherence) and a non-detected ingested marker (nonadherence), as found by the networked system. |
| VOT (AI result) | Deep Convolutional Neural Networks (DCNNs) used to extract visual features from medication intake videos, and support vector machine (SVMs) adopted as a classifier to generate prediction scores for videos, pre-trained with external datasets. Known as 3D ResNet (6). | The generated prediction score is a decimal number between 0 and 1, which can be interpreted as the probability that the video represents a participant correctly ingesting their medication. A threshold of 0.6 was used to identify adherence. |
| Mobile application (reader software) | Includes a patient and treatment supporter facing mobile app and a direct drug metabolite test. The app allows patients to report self-administration of their TB medication, track potential medication side-effects, and upload a photo of their isoniazid (INH) urine test to verify their adherence. A reader software analyzes submitted urine test images for the presence of isoniazid metabolite. Branded as 'TB-TSTs'.(7) The app also includes access to TB education, a calendar view of treatment progress, and the ability to communicate with a Treatment Supporter or other people with TB anonymously. | Reader software extracts the results area from the images and analyzes the level of color in the urine test and control strips to report detected (adherence) or not detected (nonadherence). If the drug metabolite is present the test turns a blue-purple color within 20 minutes. Refer to INH Urine Test for more details. |

#### (B) Reference Standards

| Test | Details | Adherence and nonadherence classification |
| --- | --- | --- |
| INH Urine Test | The isoniazid (INH) urine test mixes reagents with a participant's urine sample to determine the presence of drug metabolites. Branded as 'IsoScreen'. | Colour change indicates the time of last medication ingestion: Purple/blue (<24 hours ago), Green (24-48 hours ago), Yellow (>48 to 72 hours ago)<br>Refer to Table 2 for relevant time windows in each study.<br><br>*van den Boogaard et al.*(8)*:* Participants with at least one negative test were regarded as non-adherent. Tests were conducted at routine clinic visits.<br>*Thomas et al.*(3)*, Subbaraman et al.*(2)*:* Tests were conducted during an unannounced home visit.<br>*Alacapa et al.*(9)*:* Tests were conducted at a random clinic visit.<br>*Efo et al.*(4)*:* The setting in which the urine test was conducted was unclear. |
| RIF Urine Colour Test | Either the raw sample is checked for an orange colour, indicative of the presence of rifampicin (RIF) in the body, or alternatively, rifampicin is soluble in chloroform and trichloromethane so urine appears red when these are added. | Orange or red urine (adherence). Yellow urine (nonadherence).<br><br>*van den Boogaard et al.*(8)*:* Participants with at least one negative test were regarded as non-adherent. Tests were conducted at routine clinic visits.<br>*Huan et al.*(10)*:* Test was conducted at an unannounced home visit. |
| Pill Count | The participant's remaining pills were counted by a healthcare provider. | A correct (adherence) or incorrect (nonadherence) number of pills remaining. Refer to Table 2 (main text) for relevant pill counts in each study.<br>Records were assessed at provider visits during treatment.<br><br>*Scott et al.*(5)*:* Pill count was assessed at the end of TB infection treatment. |
| DOT | Participants received their TB medication during in-person clinic visits or home visits by a healthcare provider. | All doses were ingested via DOT (adherence) to evaluate the performance of the ingestible sensor technology.<br><br>*Belknap et al.*(11)*:* After two clinic visits, participants and investigators could choose to complete the remaining visits as field-DOT in the participant's home. |
| VOT (provider reported) | Manual revision of the video by 3 reviewers who included a trained student annotator, a senior computer scientist and a physician with expertise in medication adherence. The protocol was summarized into three basic rules that guided labelling videos as positive-actual medication ingestion activity, negative-no medication intake activities or ambiguous-if no pills were seen but there was a blurry image of a face. | 3 reviewers agreeing that the participant did (adherence) or did not (nonadherence) take the medication in the video.<br><br>*Sekandi et al.*(6)*:* Ambiguous videos were not retained for further analysis. |
| Mobile application (provider reported) | Includes a participant and treatment supporter facing mobile app and a direct drug metabolite test. Treatment Supporter or Research Staff analyzes submitted INH urine test images for the presence of isoniazid metabolite. | If the drug metabolite is present the test turns a blue-purple color within 20 minutes. Treatment supporter or research staff analyzes the urine test photo for a blue-purple colour (adherence) or otherwise (non-adherence). Refer to INH Urine Test for more details. |

AI artificial intelligence, DOT directly observed therapy, MEMS medication event monitoring system, TB-TSTs tuberculosis treatment support tool, VOT video observed therapy

**Table S5: Reported and calculated performance outcomes of interest in the included studies.**

| Study ID | DAT | Outcomes | | | | | | |
|---|---|---|---|---|---|---|---|---|
| | | Sensitivity | Specificity | PPV | NPV | Accuracy | AUC | Numbers of TP, FN, TN, or FP |
| Browne et al. 2019 (12) | Ingestible sensors | ✓ (green) | | | | | | ✓ (green) |
| Belknap et al. 2013 (11) | Ingestible sensors | ✓ (green) | | | | | | ✓ (green) |
| Scott et al. 2023 (5) | Digital pillboxes | ✓ (yellow) | ✓ (yellow) | ✓ (yellow) | ✓ (yellow) | ✓ (yellow) | | ✓ (green) |
| Bionghi et al. 2018§ (13) | Digital pillboxes | ✓ (green) | ✓ (green) | ✓ (yellow) | ✓ (yellow) | ✓ (yellow) | | ✓ (green) |
| Huan et al. 2012 (10) | Digital pillboxes | ✓ (green) | ✓ (green) | ✓ (yellow) | ✓ (yellow) | ✓ (yellow) | | ✓ (green) |
| van den Boogaard et al. 2011 (8) | Digital pillboxes | ✓ (green) [shaded] | ✓ (green) [shaded] | ✓ (green) [shaded] | ✓ (green) [shaded] | ✓ (green) [shaded] | [shaded] | ✓ (green) [shaded] |
| Ruslami et al. 2008 (14) | Digital pillboxes | ✓ (green) [shaded] | ✓ (green) [shaded] | ✓ (green) [shaded] | ✓ (green) [shaded] | ✓ (yellow) [shaded] | [shaded] | ✓ (green) [shaded] |
| Subbaraman et al. 2021§§ (2) | Medication sleeves with phone calls | ✓ (green) | ✓ (green) | ✓ (green) | ✓ (green) | ✓ (green) | ✓ (green) | ✓ (green) |
| Efo et al. 2021 (4) | Medication sleeves with phone calls | ✓ (yellow) | ✓ (yellow) | ✓ (green) | ✓ (green) | ✓ (yellow) | | ✓ (green) |
| Thomas et al. 2020 (3) | Medication sleeves with phone calls | ✓ (green) | ✓ (green) | ✓ (green) | ✓ (green) | ✓ (green) | | ✓ (green) |
| Alacapa et al. 2020† (9) | Medication sleeves with phone calls | ✓ (green) | ✓ (yellow) | ✓ (green) | ✓ (green) | ✓ (yellow) | | ✓ (green) |
| Sekandi et al. 2023 (6) | VOT (AI result) | ✓ (green) | ✓ (green) | ✓ (green) | | | ✓ (green) | ✓ (green) |
| Goodwin et al. 2022† (15) | Mobile application (Software result) | ✓ (green) | ✓ (green) | | | | | |

✓ (green) = reported

✓ (yellow) = calculated based on provided outcomes

☐ (shaded) = DAT was used as the reference standard of the published study. Performance outcomes of the pillbox were calculated from originally published values on sensitivity, specificity, PPV, and NPV.

AI artificial intelligence, AUC area under receiver operating characteristic curve, VOT video observed therapy,

§ Inpatient cohort

§§ Same cohort as Thomas et al. 2020

† Abstract from the World Conference on Lung Health of the International Union Against Tuberculosis and Lung Disease (The Union)

**Table S6: Performance of DATs assessed under controlled conditions against a reference standard.** For ingestible sensors, sensitivity was calculated according to positive detection accuracy (# of ingestible sensors detected/# of ingestible sensors ingested). Additionally, specificity, PPV, NPV, and accuracy were not estimable in the absence of false positives and true negatives.

| Study ID | DAT | DAT Adherence Classification | Reference Standard | N§ | TP | FP | FN | TN | Sensitivity (95% CI) | Specificity (95% CI) | PPV (95% CI) | NPV (95% CI) | Accuracy (95% CI) |
|---|---|---|---|---|---|---|---|---|---|---|---|---|---|
| Browne et al. 2019 (12) | Ingestible sensor | 1 detected dose | DOT | 77 (685) | 680 | 0 | 5 | 0 | 99% (98-100%)§§ | N/A | N/A | N/A | N/A |
| Belknap et al. 2013 (11) | Ingestible sensor | 1 detected dose | DOT | 30 (1080) | 1026 | 0 | 54 | 0 | 95% (94-96%) | N/A | N/A | N/A | N/A |
| Sekandi et al. 2023 (6) | VOT (AI result) | 1 detected dose | VOT (research team result) | 51 (497) | 385 | 41 | 20 | 51 | 95% (SD 2.6)† | 55% (SD 6.5)† | 90% (SD 1.8)† | NR | NR |
| Goodwin et al. 2022 (15) | Mobile App (Reader software result) | 1 detected dose | Mobile App (treatment supporter result) | NR (689) | NR | NR | NR | NR | 81% | 95% | NR | NR | NR |
| | | | Mobile App (research staff result) | NR (689) | NR | NR | NR | NR | 86% | 91% | NR | NR | NR |

AI artificial intelligence, CI confidence interval, DAT digital adherence technology, DOT directly observed therapy, FN false negatives, FP false positives, N number of participants, NPV negative predictive value, NR not reported and incalculable, PPV positive predictive value, TN true negatives, TP true positives, VOT video observed therapy

§ The number of videos or images assessed for performance across the participants are shown in parentheses.

§§ Confidence intervals account for clustering.

† Arithmetic mean and standard deviation were calculated based on 5-fold cross validation using 497 videos (51 participants). 364 videos (157 ambiguous or uncertain videos; 152 poor quality videos, 55 damaged videos) were excluded as there was not complete agreement of the classification across the three reviewers.

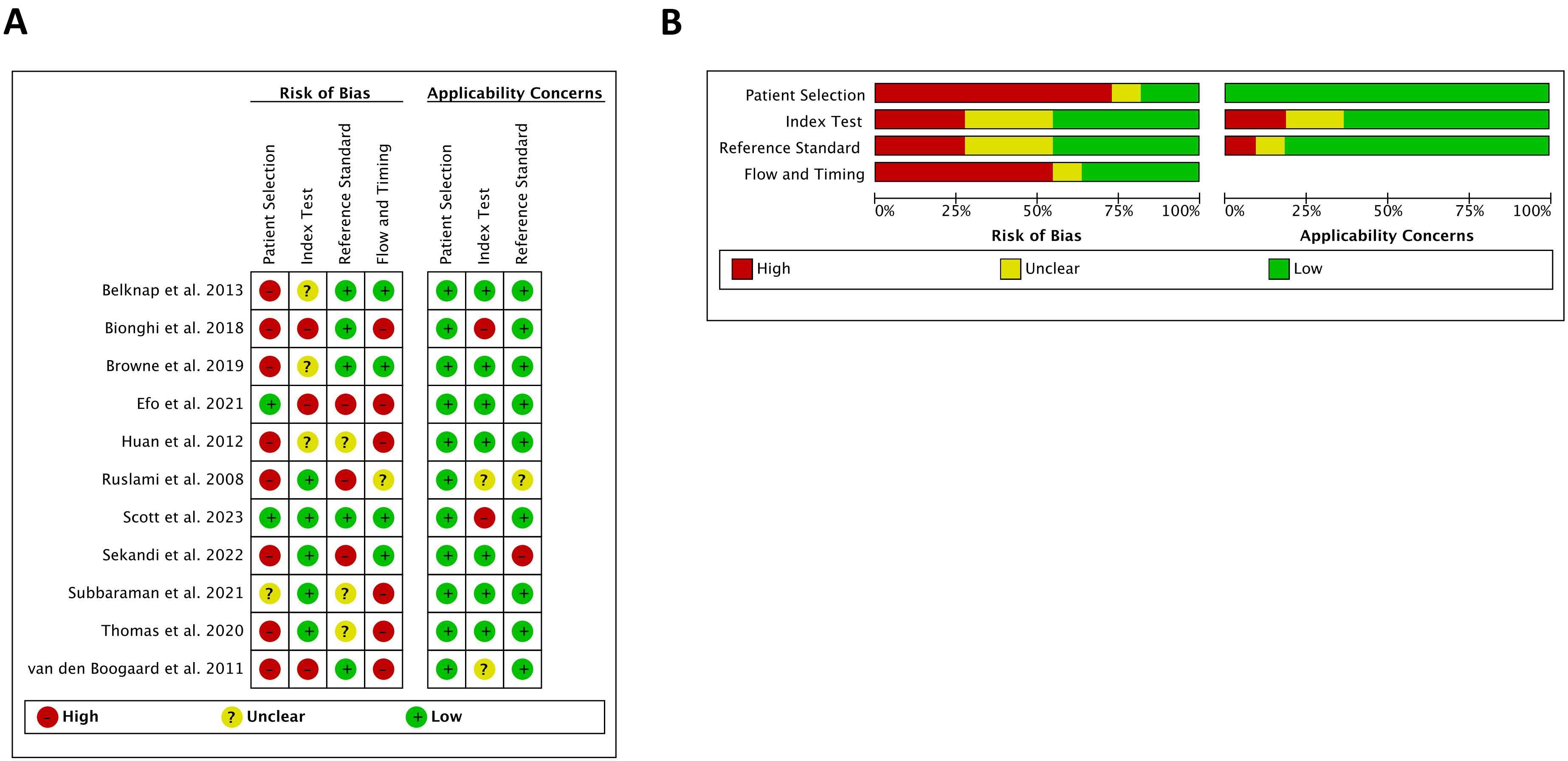


**Figure S1: Risk of bias (A) and applicability concerns (B) of included studies using the QUADAS-2 tool for diagnostic accuracy studies.** Risk of bias and applicability concerns were assessed using the Quality Assessment of Diagnostic Accuracy Studies 2 (QUADAS-2) tool for primary diagnostic accuracy studies. Alacapa et al. 2020 (9) and Goodwin et al. 2022 (15) are abstracts of the World Conference on Lung Health of the International Union Against Tuberculosis and Lung Disease (The Union) and could not be assessed for quality.

**Table S7: QUADAS-2 checklist assessment criteria for the performance of DATs for measuring TB medication adherence.**

| Question Category | QUADAS-2 Checklist Question | Alterations to QUADAS-2 Checklist Question | Assessment Criteria |
|---|---|---|---|
| **Patient Selection** | Could the selection of patients have introduced bias? | | • Any non-random sample was assessed to be a high risk of bias.<br>• Any inappropriate exclusions were assessed to be a high risk of bias. |
| | Is there concern that the included patients do not match the review question | | • Participants with available demographic information were assessed to be of low concern. |
| **Index Test** | Could the conduct or interpretation of the index test have introduced bias? | Could the conduct or interpretation of the **DAT intervention** have introduced bias? | • If other medicine was administered at the same time of DAT use or behaviour changed as a result of anticipating adherence evaluations, there was a high risk of bias.<br>• If there were DAT accessibility issues for clients there was a high risk of bias.<br>• If it was not clear how/when the DAT reports were assessed relative to the reference standard there was an unclear risk of bias. |
| | Is there concern that the index test, its conduct, or interpretation differ from the review question? | Is there concern that the **DAT intervention**, its conduct, or interpretation differ from OUR review question? | • If performance was assessed in an inpatient cohort, there was a high applicability concern.<br>• If the DAT was not initially assessed as the index test, there were unclear or high applicability concerns based on the provided performance results. |
| **Reference Standard** | Could the reference standard, its conduct, or its interpretation have introduced bias? | | • If it was not clear how/when the DAT reports were assessed relative to the reference standard there was an unclear risk of bias.<br>• If it was not clear how/when the DAT reports were assessed relative to the reference standard AND there was missing information on details of the reference standard there was a high risk of bias.<br>• If the reference standard had the ability to misclassify participants to a certain degree, there was an unclear risk of bias. |
| | Is there concern that the target condition as defined by the reference standard does not match the review question? | | • If information on the performance of the reference standard was provided and misclassification was discussed, there was an unclear applicability concern.<br>• If there was not enough detail provided on the reference standards to assess this question, there was an unclear applicability concern.<br>• If the reference standard was conducted by individuals that would not typically assess adherence for those with TB, there was a high applicability concern. |
| **Flow and Timing** | Could the patient flow have introduced bias? | | • If performance was assessed at only one point during the course of treatment to depict adherence, there was a high risk of bias.<br>• If a large number of participants were removed from the analysis/performance assessment, there was a high risk of bias.<br>• If it was not described when participants underwent each test, there was an unclear risk of bias. |

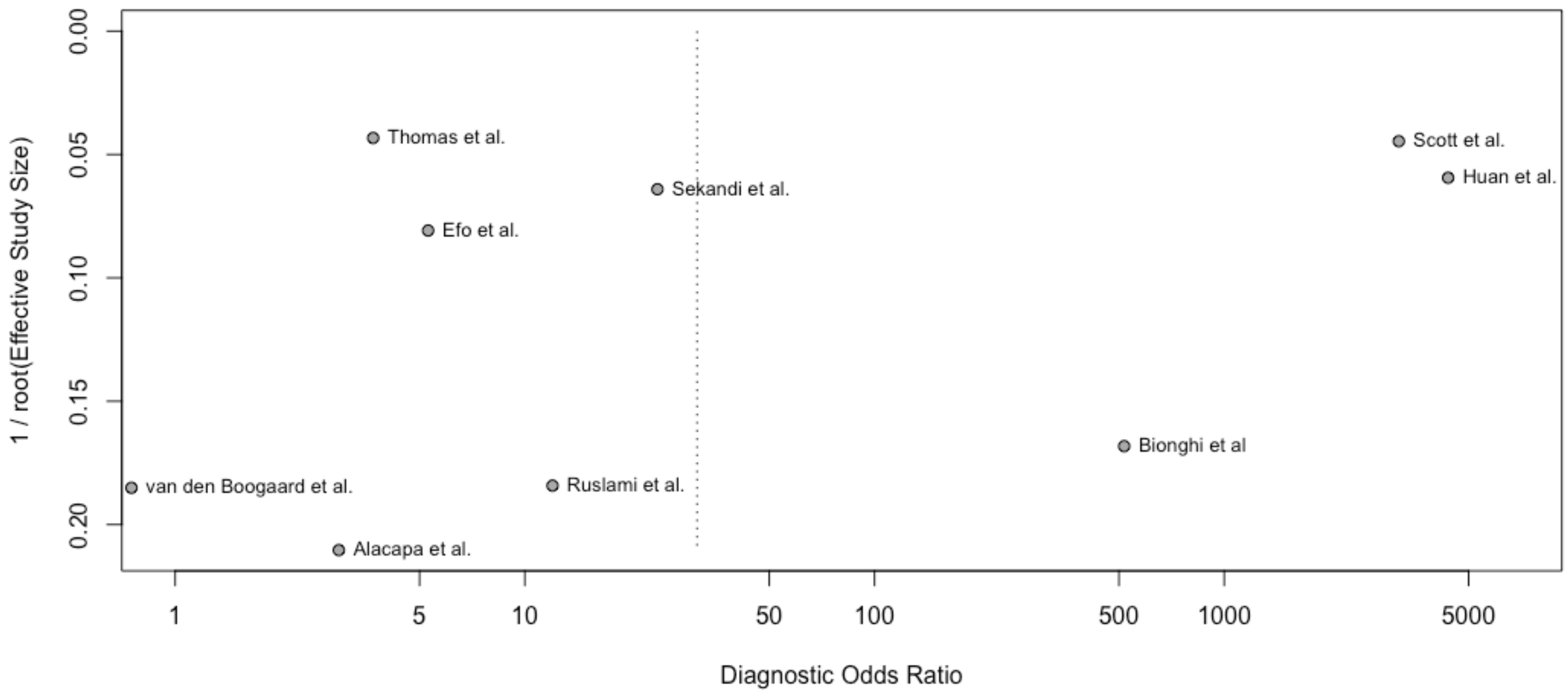


**Figure S2: Funnel plot of included studies for the qualitative assessment of publication bias.** Publication bias was assessed using Deek's test for diagnostic odds ratios. DOR is the odds of a positive dose record by the DAT in those who were adherent (based on the reference standard) relative to the odds of a positive dose record in those who were not adherent while ESS is a function of the number of adherent ($n_1$) and non-adherent ($n_2$) participants ($(4n_1 * n_2)/(n_1 + n_2)$).(16) The results shown in the forest plots for each DAT, or the primary result in the data table, was used to assess publication bias. Subbaraman et al. (2) used the same primary cohort as Thomas et al. (3) and was not included in the publication bias assessment. Belknap et al. (11) and Browne et al. (12) only assessed sensitivity of ingestible sensors where no diagnostic odds ratio could be calculated and are not included in this analysis. Goodwin et al. (15) did not provide enough information to assess publication bias. Deek's test uses a linear regression of lnDOR with 1/Effective Sample Size$^{1/2}$ weighted by the effective sample size.

**Table S8: The quality of evidence for the performance of ingestible sensors for measuring TB medication adherence.** Assessed using criteria of the Grading of Recommendations, Assessment, Development and Evaluation (GRADE) approach for diagnostic tests and strategies**.** The quality of evidence for true negatives and false positives could not be assessed due to only true positives and false negatives provided as positive detection accuracy being provided in each study (# of ingestible sensors detected/# of ingestible sensors ingested).

**Should Ingestible sensors be used to report on medication adherence in those with TB disease?**

**Patient or population**: those with TB disease
**Setting**: Outpatient TB treatment
**New test**: Ingestible sensors | **Cut-off value**: 1 detected dose (adherence)
**Reference test**: Directly observed therapy (DOT) | **Threshold**: 1 ingested dose in person (adherence)
**Range of sensitivities**: 0.95 to 0.99 | **Range of specificities**: -- to --

| Test result | Number of results per 1,000 patients tested (95% CI) | | Number of participants (studies) | Certainty of the Evidence (GRADE) |
|---|---|---|---|---|
| | **Prevalence = 80%** Typically seen in | **Prevalence = 95%** Typically seen in | | |
| **True positives** | 760 to 792 | 903 to 941 | 107 (2) | ⊕⊕⊕◯ **Moderate**[a,b] |
| **False negatives** | 8 to 40 | 9 to 47 | | |
| **True negatives** | 0 to 0 | 0 to 0 | (0) | - |
| **False positives** | 200 to 200 | 50 to 50 | | |

**CI:** confidence interval

#### *Explanations*

a. Specificity was unable to be assessed due to how the performance of the ingestible sensor technology was assessed compared to directly observed therapy where all doses were known and taken.
b. Publication bias could not be assessed due to only sensitivity being estimated and not possible to assess using Deek's test for diagnostic odds ratios. Therefore, test performance should be interpreted with caution.

**Table S9: The quality of evidence for the performance of medication sleeves for measuring TB medication adherence.** Assessed using criteria of the Grading of Recommendations, Assessment, Development and Evaluation (GRADE) approach for diagnostic tests and strategies. The quality of evidence for Subbaraman et al. (2) was not assessed as it has the same cohort as Thomas et al. (3). The result present in the forest plots of medication sleeves with phone calls (Figure 2b, main text) was used to assess the quality of evidence.

**Should Medication sleeves with phone calls be used to report on medication adherence in those with TB disease?**

**Patient or population**: those with TB disease
**Setting**: Outpatient TB treatment
**New test**: Medication sleeves with phone calls (e.g., 99DOTS) | **Cut-off value**: 1 dose reported in the previous 48 hours (adherence)
**Reference test**: Isoniazid Urine Metabolite Test | **Threshold**: Purple/blue or green colour change (dose taken in previous 48h – adherence)
**Range of sensitivities**: 0.70 to 0.94 | **Range of specificities**: 0.01 to 0.61

| Test result | Number of results per 1,000 patients tested (95% CI) | | Number of participants (studies) | Certainty of the Evidence (GRADE) |
|---|---|---|---|---|
| | **Prevalence = 80%** Typically seen in | **Prevalence = 95%** Typically seen in | | |
| **True positives** | 560 to 752 | 665 to 893 | 805 (3) | ⨁◯◯◯ **Very low**[a,b] |
| **False negatives** | 48 to 240 | 57 to 285 | | |
| **True negatives** | 2 to 122 | 1 to 31 | 92 (3) | ⨁◯◯◯ **Very low**[a,c,d] |
| **False positives** | 78 to 198 | 19 to 49 | | |

**CI:** confidence interval

#### *Explanations*

a. One article was an abstract, wherein the risk of bias could not be properly assessed due to a lack of information
b. Sensitivity was variable across articles, with non-overlapping 95% confidence intervals
c. Specificity had variable estimates but overlapping 95% confidence intervals. However, due to the strong variability in estimates, it was deemed serious.
d. There are very wide 95% confidence intervals for specificity, with one study having a 95% CI range from 0% to 84%.

**Table S10: The quality of evidence for the performance of digital pillboxes for measuring TB medication adherence.** Assessed using criteria of the Grading of Recommendations, Assessment, Development and Evaluation (GRADE) approach for diagnostic tests and strategies. Bionghi et al. (13) was not included because the article reported performance for the total doses of the sample rather than total participants and it used an inpatient cohort. Huan et al. (10) was not included because it uses a different reference standard (RIF urine colour test). Scott et al. (5) was not included because it assessed the performance of digital pillboxes in those with TB infection. The result present in the forest plots of digital pillboxes (Figure 2c, main text) was used to assess the quality of evidence.

**Should Digital pillboxes be used to report on medication adherence in those with TB disease?**

**Patient or population**: those with TB disease
**Setting**: Outpatient TB treatment
**New test**: Digital pillboxes (e.g., Wisepill, MEMS) | **Cut-off value**: 100% of doses (adherence)
**Reference test**: Pill count | **Threshold**: 100% of doses (adherence)
**Range of sensitivities**:0.25 to 0.75 | **Range of specificities**: 0.69 to 0.80

| Test r esult | Number of results per 1,000 patients tested (95% CI) | | Number of participants (studies) | Certainty of the Evidence (GRADE) |
|---|---|---|---|---|
| | Prevalence = 80% Typically seen in | Prevalence = 95% Typically seen in | | |
| **True positives** | 200 to 600 | 238 to 712 | 44 (2) | ⊕◯◯◯ **Very low**[a,b,c,d] |
| **False negatives** | 200 to 600 | 238 to 712 | | |
| **True negatives** | 138 to 160 | 35 to 40 | 23 (2) | ⊕◯◯◯ **Very low**[a,c,d,e] |
| **False positives** | 40 to 62 | 10 to 15 | | |

**CI:** confidence interval

#### *Explanations*

a. Two reports originally assessed the digital pillbox as the reference standard and we back-calculated to determine the sensitivity and specificity for the digital pillbox as the index test.
b. Sensitivity has highly variable across studies, with non-overlapping confidence intervals
c. Due to the back-calculation of the performance of digital pillboxes, there are applicability concerns when answering the research question.
d. There were very wide 95% confidence intervals for two of the articles for serious imprecision.
e. Specificity had similar estimates across all studies and overlapping confidence intervals across two of the studies.

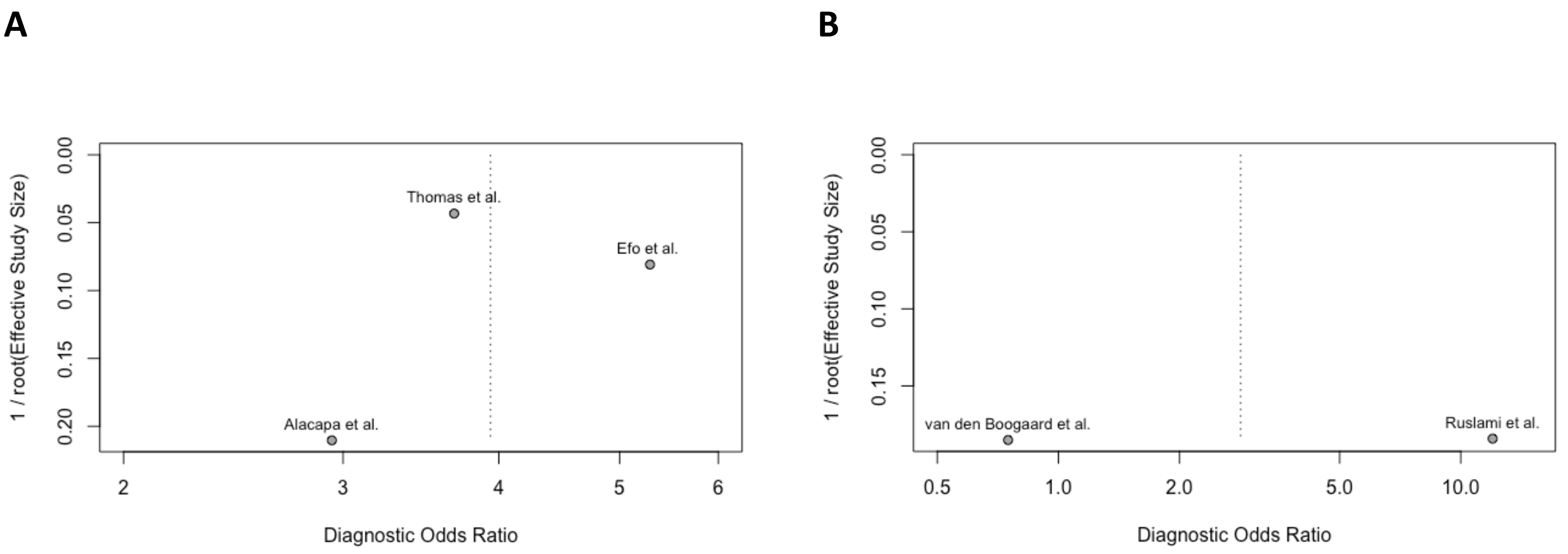


**Figure S3: Publication bias using Deek's test of articles reporting on each DAT.** (A) Publication bias for medication sleeves with phone calls and (B) Publication bias for digital pillboxes as analyzed for the GRADE assessment. **(A)** Subbaraman et al. (2) uses the same primary cohort as Thomas et al. (3) and was not included in the GRADE assessment. **(B)** Bionghi et al. (13) was not included because the article reported on performance for the total doses of the sample rather than total participants and it used an inpatient cohort. Huan et al. (10) was not included because it uses a different reference standard (RIF urine colour test). Scott et al. (5) was not included because it assessed the performance of digital pillboxes in those with TB infection. For ingestible sensors, Belknap et al. (11) and Browne et al. (12) only assessed sensitivity wherein no diagnostic odds ratio could be calculated and therefore, no publication bias could be assessed. Deek's test uses a linear regression of lnDOR with 1/Effective Sample Size$^{1/2}$ weighted by the effective sample size.

10. HUAN Shi-tong LIU Xiao-qiu, OU Xi-chao, JIANG Shi-wen, ZHAO Yan-lin, ZHANG Zhi-ying, ZHAN Si-yan CR. Operational feasibility of medication monitors in monitoring treatment adherence among TB patients [Internet]. Vol. 34, Chinese Journal of Antituberculosis. p. 419–24. Available from: http://www.zgflzz.cn

11. Belknap R, Weis S, Brookens A, Au-Yeung KY, Moon G, DiCarlo L, et al. Feasibility of an Ingestible Sensor-Based System for Monitoring Adherence to Tuberculosis Therapy. PLoS One. 2013;8(1):8–

12.

12. Browne SH, Umlauf A, Tucker AJ, Low J, Moser K, Garcia JG, et al. Wirelessly observed therapy compared to directly observed therapy to confirm and support tuberculosis treatment adherence: A randomized controlled trial. PLoS Med. 2019;16(10):1–19.

13. Bionghi N, Daftary A, Maharaj B, Msibi Z, Amico KR, Friedland G, et al. Pilot evaluation of a second-generation electronic pill box for adherence to Bedaquiline and antiretroviral therapy in drug-resistant TB/HIV co-infected patients in KwaZulu-Natal, South Africa. BMC Infect Dis. 2018;18(1):1–9.

14. Ruslami R, Van Crevel R, Van De Berge E, Alisjahbana B, Aarnoutse RE. A step-wise approach to find a valid and feasible method to detect non-adherence to tuberculosis drugs. Southeast Asian J Trop Med Public Health. 2008;39(6):1083–7.

15. Goodwin K, Liao Z, Iribarren S. Assessing and refining image analysis software to automate reading of objective home-based TB adherence tests. World Conf Lung Heal Int Union Against Tuberc Lung Dis (The Union). 2022;26.

16. Van Enst WA, Ochodo E, Scholten RJ, Hooft L, Leeflang MM. Investigation of publication bias in meta-analyses of diagnostic test accuracy: A meta-epidemiological study. BMC Med Res Methodol. 2014;14(1):1–11.
